# Effect of Sevelamer versus *Bifidobacterium longum* on Insulin Sensitivity in Subjects with Obesity

**DOI:** 10.1101/2025.06.23.25327556

**Authors:** Eric Baeuerle, Manpreet K Semwal, Jia Nie, Ning Zhang, Hanyu Liang, Vinutha Ganapathy, Nattapol Sathavarodom, Roman Fernandez, Chen-Pin Wang, Sara Espinoza, Qunfeng Dong, Zhen Yang, Aleksandar Kostic, Nicolas Musi

## Abstract

**Objective:** To test whether interventions with putative lipopolysaccharide (LPS)-lowering effects including sevelamer and a synbiotic (Bifidobacterium longum+oligofructose) improve insulin sensitivity in subjects with obesity.

**Methods:** We randomized 22 lean and 28 human subjects with obesity to receive sevelamer, synbiotic, or placebo orally for 4 weeks. Peripheral insulin sensitivity was measured with an insulin clamp. Plasma cholesterol, endotoxemia markers, intestinal permeability, stool bacterial taxonomic content and plasma metabolites also were assessed.

**Results:** At baseline, subjects with obesity had lower insulin sensitivity and elevated plasma LDL-C concentrations. Sevelamer improved insulin sensitivity and lowered LDL-C in these subjects. Intestinal enrichment of *Bifidobacterium longum* had no effect on insulin sensitivity or lipids in either group. Markers of endotoxemia and intestinal permeability did not change with any intervention. Plasma metabolomics revealed that sevelamer increases several metabolites, many of them previously linked with positive changes in glucose and lipid metabolism, including biliary acids, amino acids (citrulline, betaine), NAD+ precursors (trigonelline) and xenobiotics (genistein, umbelliferone).

**Conclusion:** Sevelamer improves insulin sensitivity and LDL-C in subjects with obesity, but these effects are independent of changes in circulating LPS. Sevelamer increases the levels of multiple metabolites that collectively may mediate improvements in glucose homeostasis and lipids caused by this drug.

## INTRODUCTION

Insulin resistance in peripheral tissues (i.e., skeletal muscle) is one of the earliest and most significant abnormalities in obesity and type 2 diabetes mellitus. However, the etiology of insulin resistance is not fully understood. Increasing evidence suggests that the gut microbiome plays a role in the pathophysiology of insulin resistance, obesity, and type 2 diabetes, and may represent a therapeutic target against them. For example, some studies indicate that bacteria with putative health-promoting properties such as bacteria of the phylum Bacteroidetes and the genus *Bifidobacterium* are decreased in subjects with obesity and type 2 diabetes (1). Moreover, transplantation of gut microbiota from high fat fed mice (as opposed to low fat fed mice) into germ-free mice increases adiposity and promotes metabolic dysfunction (2).

The gut microbiome is hypothesized to modulate glucose homeostasis via different mechanisms, including the production of substances which alter intestinal wall integrity, energy extraction from food, energy expenditure, and appetite (reviewed in (3)). In particular, it is postulated that the microbiome affects glucose metabolism via a mechanism that involves translocation of lipopolysaccharides (LPS) from the gut lumen through the intestinal barrier and into the circulation; this phenomenon has been termed metabolic endotoxemia (4). LPS is a component of the outer membrane of bacteria cell walls, which induces an inflammatory response by activating toll-like receptor (TLR)-4. Subjects with obesity and type 2 diabetes have increased LPS concentrations in the circulation (5, 6) and ingestion of high fat meals increases circulating LPS level in humans (7). Conversely, modulation of the intestinal microbiota composition with prebiotics and antibiotics reduces plasma LPS concentration and improves glucose intolerance in insulin resistant mice (8, 9).

Despite the accumulating evidence suggesting that gut microbiota and metabolic endotoxemia play an important role in the pathogenesis of insulin resistance, obesity, and type 2 diabetes, the relevance of the intestinal microbiome and metabolic endotoxemia in human metabolic disease remains unclear. In this study, we hypothesized that two interventions with putative LPS-lowering effects, the non-absorbable polymer sevelamer (9), and the synbiotic (a combination of a prebiotic and a probiotic) made of *Bifidobacterium longum* plus oligofructose (10), would improve metabolic outcomes in subjects with obesity.

## METHODS

### Subjects

Subjects were enrolled via local advertisement in the San Antonio, TX, area. All study procedures took place in the Bartter Research Unit at the Audie L. Murphy VA Medical Center. The study was approved by the IRB of the University of Texas Health Science Center at San Antonio and registered in Clinicatrials.gov (NCT02127125). All subjects gave written informed consent.

The study was open to participants of both sexes and all races and ethnicities and was conducted from April 2014 to August 2018. Subjects did not take drugs known to impact glucose and lipid metabolism, probiotics, prebiotics, and antibiotics within 3 months. All subjects were normal-glucose tolerant, generally healthy (no history of inflammatory, hematologic, pulmonary, cardiovascular, hepatic, kidney, gastrointestinal or neurologic disease), with a stable body weight (no more than ±2% change in body weight over the last 3 months) and sedentary (≤2 sessions of exercise/week for previous 6 months). Two groups of subjects were recruited based on BMI. For the purpose of this study, lean category was defined as a BMI <26 kg/m^2^ and obese category as BMI = 30 to 37 kg/m^2^.

### Oral glucose tolerance test

To establish normal glucose tolerance, subjects underwent a standard 2-h oral glucose tolerance test after ingestion of 75 g dextrose in the fasted state.

### Study intervention

The study flowchart is shown in Figure 1. Subjects were randomized to one of three interventions (sevelamer, synbiotic, or placebo) in a double-blind fashion; block randomization was done by a research pharmacist. Other research personnel did not have access to group assignment. Subjects in the sevelamer group received 6 g/day sevelamer-HCl (Amgen, Cambridge, MA) plus 4.4 g/day maltodextrin (MuscleFeast LLC, Nashport, OH); subjects in the synbiotic group received 4×10^10^ *Bifidobacterium longum* CFU/day (Lallemand Co, Montreal, CA) plus 5 g/day oligofructose (both from Lallemand Co, Montreal, CA); and subjects in the placebo group received 6 g/day maltodextrin. Subjects received all study drug from a blinded research nurse in three divided doses as powder in packets, instructed to mix the powder with water, and ingest the mixture orally for 28 days. Adherence was assessed by counting unused study medication.

**Figure 1.**
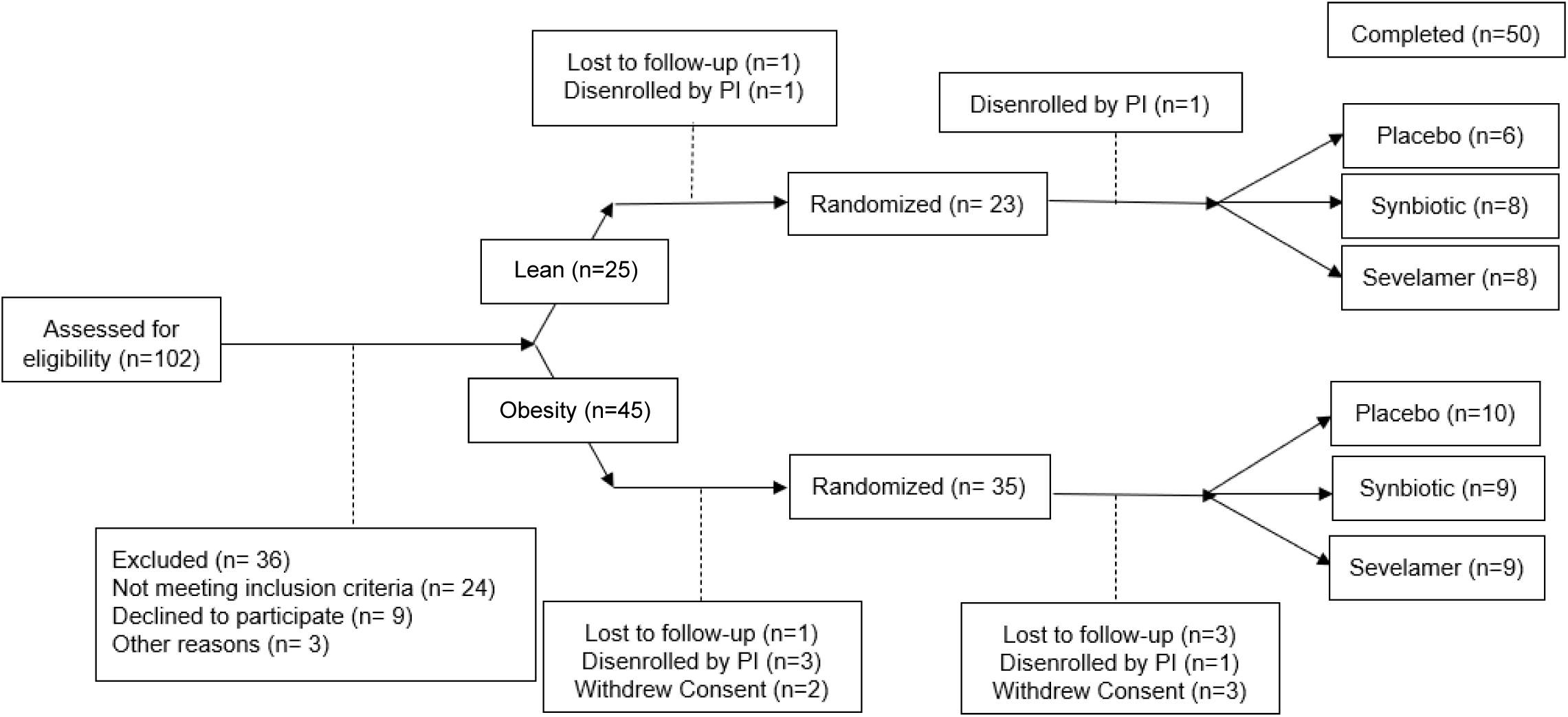
Clinical Flow of Subjects in the Microbiome Study. 102 subjects were consented and assessed for eligibility. Reasons for subjects ending participation in the study are documented at assignment to group, pre-randomization, and post-randomization prior to completing study. 7 subjects were disenrolled due to usage of antibiotics for illness unrelated to the study.

### Intestinal permeability assay

This assay was conducted because an increased lactulose:mannitol ratio indicates higher intestinal permeability (11). Subjects ingested a solution containing 5 g lactulose and 2 g mannitol in the fasted state. Lactulose and mannitol concentration were measured by colorimetric assay using an EnzyChrom Intestinal Permeability Assay Kit (BioAssay Systems, Hayward, CA) in urine at baseline and 2, 4, and 6 h after ingestion.

### Insulin clamping

Peripheral insulin sensitivity was measured with a 60 mU/m^2^.min hyperinsulinemic euglycemic clamp (12). Clamps started at ∼7-8 AM, in the fasting state. Peripheral insulin sensitivity is reported both as the M value and M value divided by plasma insulin levels (M/I) during the last 30 min of the clamp.

### Laboratory analyses

Plasma glucose was measured using an Analox analyzer (Lunenburg, MA) and hemoglobin A1c using a DCA2000 analyzer (Bayer, Tarrytown, NY). Plasma insulin was measured by radioimmunoassay (Diagnostic Products, Los Angeles, CA). Homeostatic model assessment-insulin resistance (HOMA-IR) was calculated as (FPI x FPI)/22.5. Plasma LPS levels were determined using a Limulus Amoebocyte Lysate (LAL) assay (Lonza, Walkersville, MD). Plasma IL-6 (R&D Systems), TNFα (R&D Systems), FGF-19 (R&D Systems) and zonulin (Human Haptoglobin Immunoassay, R&D Systems, Minneapolis, MN) were measured by ELISA. Lipid profiling in plasma was done by the clinical laboratory of the Audie Murphy VA Hospital.

### Microbiome sample collection and processing

Stool samples were collected by participants at home in sterile plastic containers, refrigerated, and frozen at -80°C within 1 day of collection. Bacterial DNA extraction was performed using ZymoBIOMICS DNA Kit (Zymo Research, Irvine, CA). Library preparation was completed according to previous protocols (13). Genomic sequencing was performed by GeneWIZ (South Plainfield, NJ using an Illumina HiSeq (San Diego, CA) using 2×150bp metagenomic sequencing. Samples were pooled at 16 samples per lane at a depth of 350 million reads per lane. Sequencing primers and Indexing primers were Standard Illumina.

### Metabolomics

Plasma samples collected from participants in the lean and obese groups before and after placebo and sevelamer administration were evaluated by Metabolon Inc (Durham, NC) for metabolite concentrations via liquid chromatography-mass spectrometry.

### Statistical analysis

All statistical analyses were performed on datasets by researchers blinded to intervention. The primary outcome was change in peripheral insulin sensitivity. Comparisons of clinical and metabolic variables between lean and obese groups at baseline were made using unpaired t-test or Mann-Whitney test depending on normality of data. Comparisons between baseline and post-intervention measures within a group were assessed by paired t-test. Significance for paired t-tests is set at p=0.05. Placebo controlled intervention effects are analyzed using generalized estimating equations. Significance for generalized estimating equations is set at p=0.025 to correct for multiple comparisons to evaluate randomization of intervention groups. Metabolomics data were analyzed using ANOVA contrasts.

### Gut microbiome analysis method

Raw metagenomic sequences were used as inputs to run MetaPhlAn (version 2.7.7) with default settings in order to identify species abundances in each sample. Merge_metaphlan_tables.py (which is included in MetaPhlAn) was used to create a species abundance matrix for all the samples. Phyla abundance comparison was performed in baseline subjects using Mann-Whitney test. Comparisons between baseline and post-intervention phyla abundance were assessed using Wilcoxon matched-pairs signed rank test. Significance was set at α < 0.05.

### Microbiome diversity and enrichment analysis

Species abundance matrices were assessed using the R program “vegan” (14). Species accumulation tables were developed using the function “specaccum.” Species alpha diversity (within-subject diversity) was assessed using the “diversity” function to test for Shannon-Weaver Index, Simpson Index, and Inverse Simpson Index. Evenness was assessed using Pielou’s evenness index, derived from the Shannon-Weaver Index divided by the log transformed number of species. Diversity index comparison was performed in baseline subjects using Mann-Whitney test. Comparisons between baseline and post-intervention diversity and evenness were assessed using Wilcoxon matched-pairs signed rank test. Beta diversity (between-subjects diversity) was assessed in all untransformed species level abundance analysis using the “vegdist” function in “vegan” to obtain a Bray-Curtis distance dissimilarity matrix between samples. Beta diversity in log-transformed species data and all pathway abundance analysis were assessed using “vegdist” to obtain Euclidean distance between samples. Linear Discriminate Analysis (LDA) was performed using LefSe (15), available at http://huttenhower.sph.harvard.edu/galaxy/. Significance was set at an LDA score of ±2 (Log10).

## RESULTS

### Human participant characteristics

Participant characteristics are shown in Table S1. 22 lean and 28 subjects with obesity completed the study (Figure 1). Age and sex distribution was similar between lean and obesity groups.

### Baseline glucose and cholesterol metabolism parameters

Subjects with obesity had elevated fasting plasma glucose, fasting plasma insulin, and HOMA-IR index compared with lean subjects (Figures 2A-2C). Accordingly, peripheral insulin sensitivity (M value and M/I) measured with a euglycemic hyperinsulinemic clamp was reduced in subjects with obesity (Figures 2D and 2E). In line with these findings, LDL cholesterol and triglyceride levels were elevated (Figures 2G and 2I) and HDL cholesterol was reduced in the obesity group (Figure 2H).

**Figure 2.**
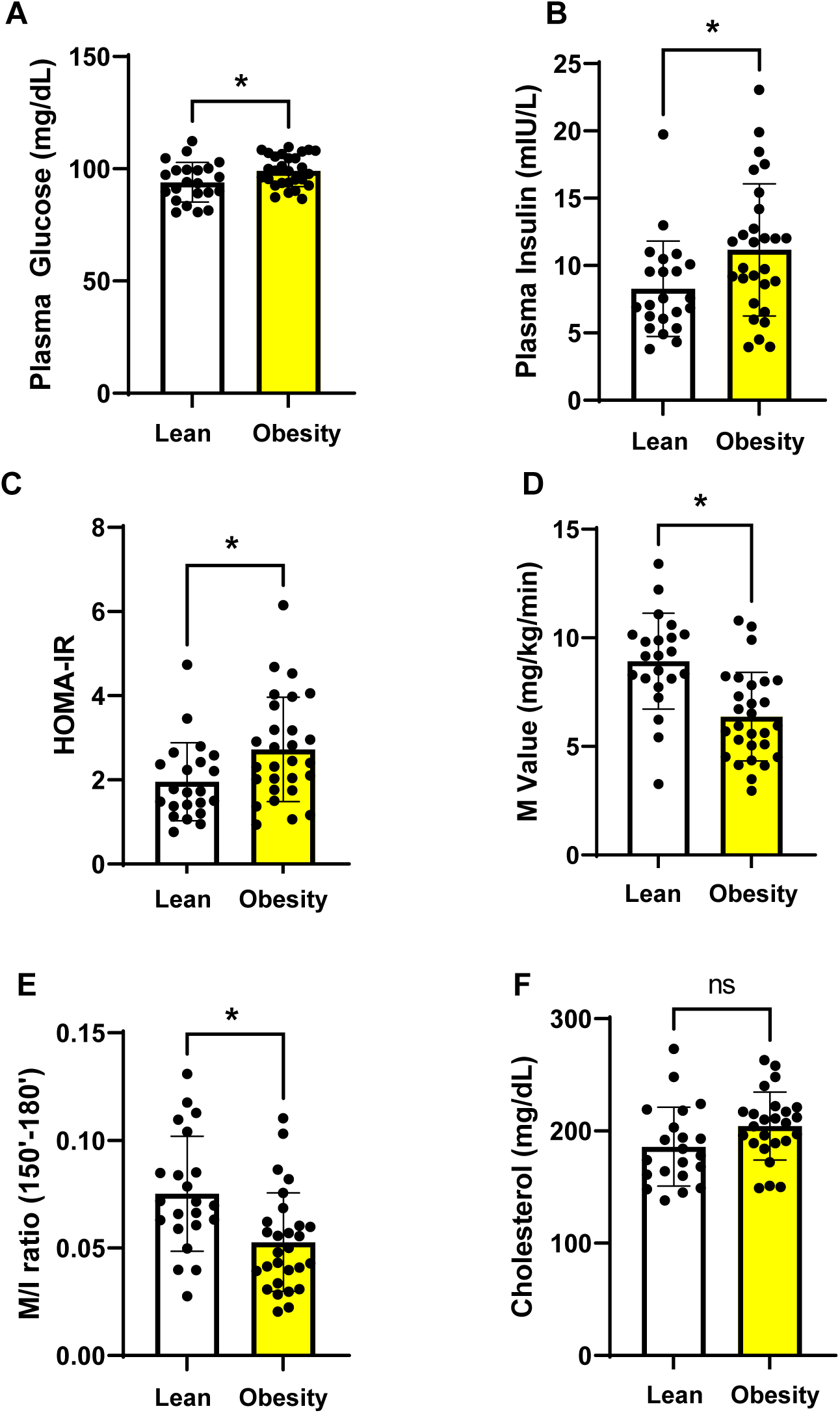
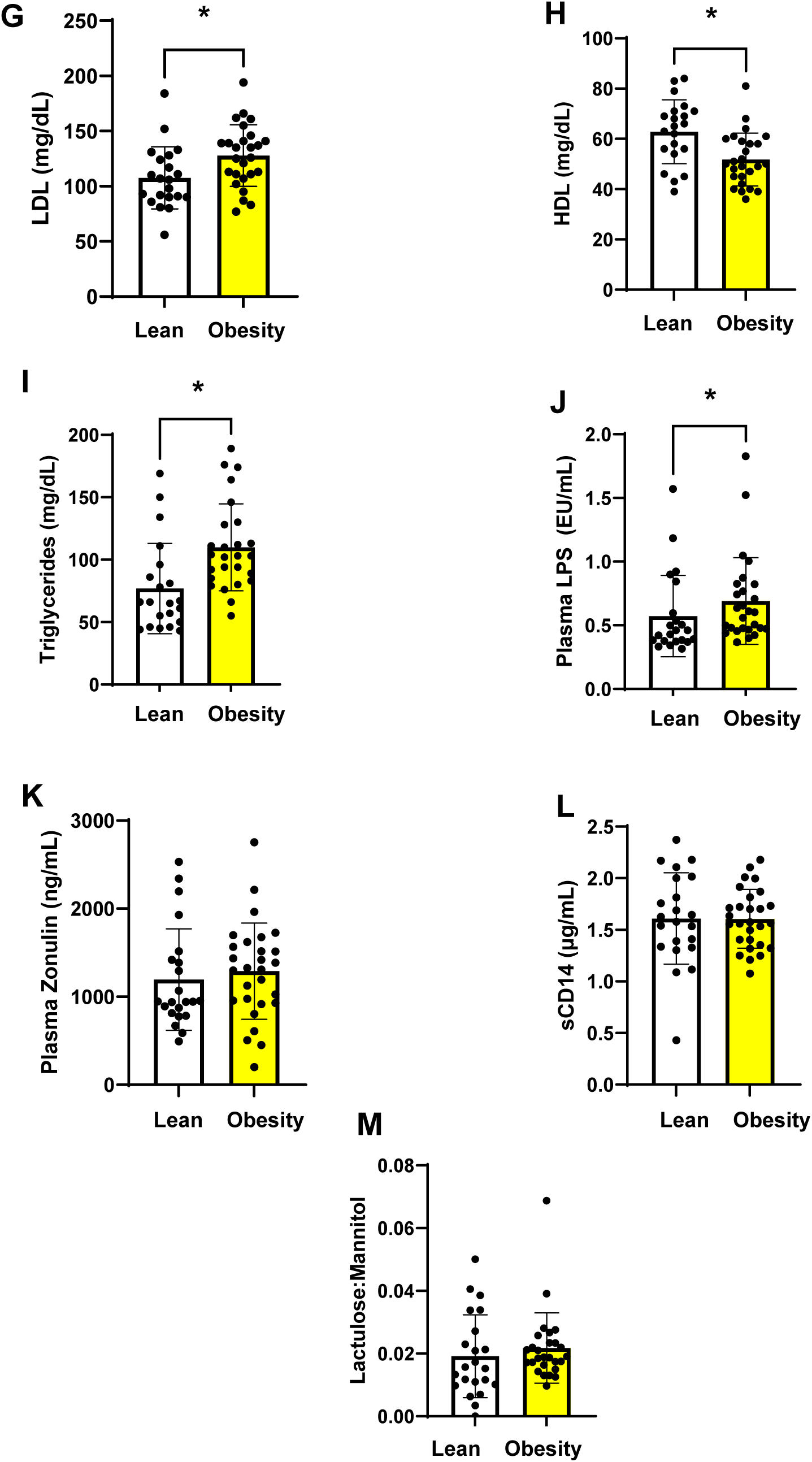
Baseline glucose metabolism outcomes, lipid profile of metabolic endotoxemia and intestinal permeability measures in lean vs obesity groups. (A) Plasma glucose, (B) insulin, (C) HOMA-IR, (D) M value, (E) M/I, (F) total cholesterol, (G) LDL-cholesterol, (H) HDL-cholesterol, (I) triglycerides, (J) LPS, (K) zonulin, (L) sCD14, and (M) lactulose:mannitol. *p<0.05 by t-test. Data are means ± SEM.

### Baseline endotoxemia and intestinal permeability markers

Subjects in the obesity group had elevated plasma LPS levels (Figure 2J). Intestinal permeability markers including plasma zonulin concentration, serum soluble (s) CD14 levels, and lactulose:mannitol ratio were similar in lean and obesity groups (Figures 2K-2M).

### Effects of sevelamer and synbiotic

Sevelamer, synbiotic, and the placebo were well tolerated and there was no difference in adverse events between groups (Table S2). In lean subjects, neither sevelamer or the synbiotic significantly affected body weight, glucose metabolism parameters, lipids, markers of endotoxemia or measures of intestinal permeability (Figures S1 – S3).

In subjects with obesity, sevelamer and synbiotic treatment were associated with a small increase in mean body weight from baseline, although there were no differences between treatment groups (Figure 3A). Sevelamer reduced plasma glucose concentrations by 7% from baseline (Figure 3B) without differences between treatment groups. Plasma insulin concentrations and HOMA-IR did not change with sevelamer or synbiotic (Figures 3C and 3D). Notably, sevelamer treatment led to a 36% increase in peripheral insulin sensitivity (M) compared with placebo and a 28% increase in M/I from baseline (Figures 3E and 3F). Synbiotic administration did not affect M and was associated with a small decrease in M/I within the treatment group, but not between treatment groups. Sevelamer reduced plasma LDL-C by 13% from baseline and this effect was significantly different versus placebo (Figure 3H). Plasma HDL-C decreased by 11% from baseline but there were no treatment group differences in the concentration of this lipoprotein (Figure 3I). Plasma triglycerides did not change in any of the treatment groups (Figure 3J).

**Figure 3.**
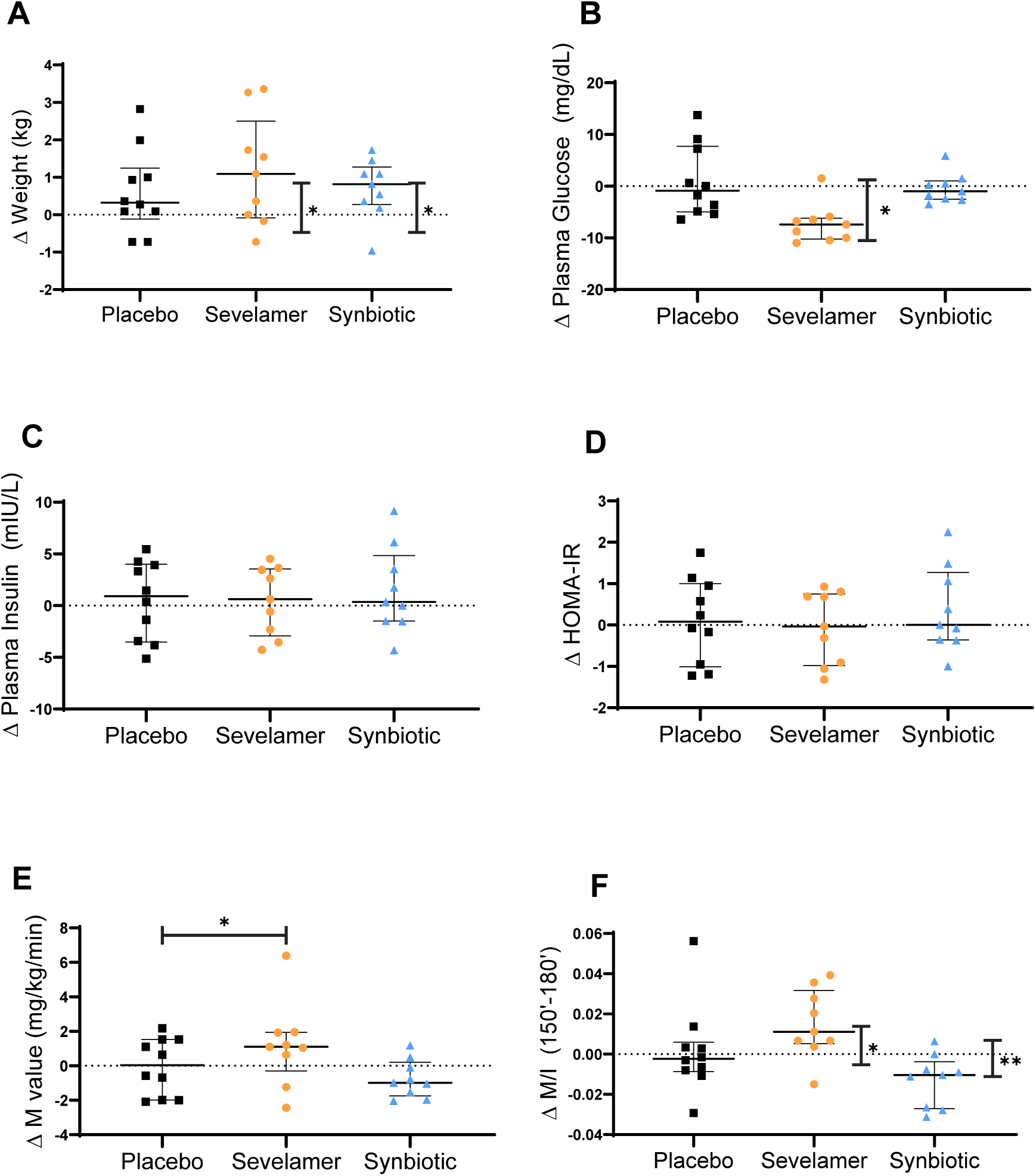
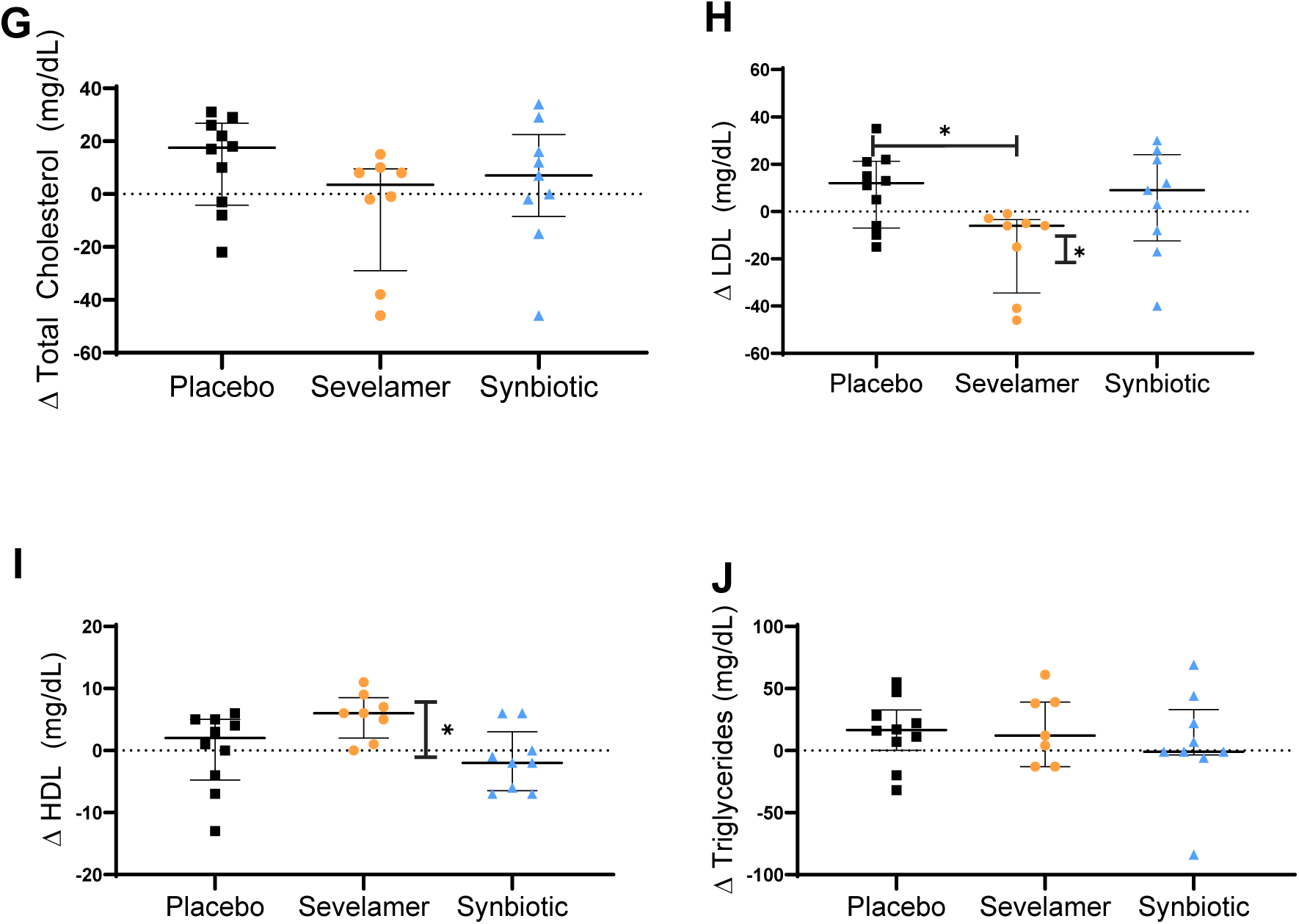
Effect of placebo, sevelamer and synbiotic on glucose metabolism and serum lipids in the obesity group. (A) Body weight, (B) plasma glucose, (C) insulin, (D) HOMA-IR, (E) M value, (F) M/I, (G) total cholesterol, (H) LDL-cholesterol, (I) HDL-cholesterol and (J) triglycerides. Within-intervention effects were analyzed using paired t-test; *p<0.05. Between intervention effects analyzed using generalized estimating equations; *p<0.025. Data are means ± SEM.

### Gut microbiome analysis

A total of 49 baseline stool samples (21 from lean and 28 subjects with obesity) yielded high quality DNA and were analyzed via metagenomic sequencing as described (13). Comparison of the four highest abundant phyla (Firmicutes, Bacteroidetes, Actinobacteria, or Veruccomicrobia) between lean and obese groups at baseline did not show significant differences in relative abundance (Figures 4A-4E). Species-level alpha diversity indices (within-subjects) were determined using ecologically derived measures. Each index is dependent on the proportional abundance of each individual species within the sample. Subjects with obesity had reduced diversity compared to lean subjects at baseline in both the Shannon-Weaver Index (p=0.05) and the Simpson and Inverse Simpson Indices (p=0.028) (Figures 4F-4G). Using Pielou’s evenness index, which identifies the relative equality of abundance of the species represented within a sample, we also found that the microbiome of obese group is less even when compared with lean subjects (p=0.047) (Figure 4H). Linear discriminate analysis (LDA) was done to identify bacteria preferentially enriched in either lean or obese groups. Leanness-associated bacteria included *Bifidobacterium longum*, *Bacteroides eggerthii,* and other previously identified species (Figure 4I), while obesity-associated bacteria included multiple bacteria from the *Megamonas* genus as well as less commonly identified *Paraprevotella* and *Phascolarctobacterium* genus bacteria.

**Figure 4.**
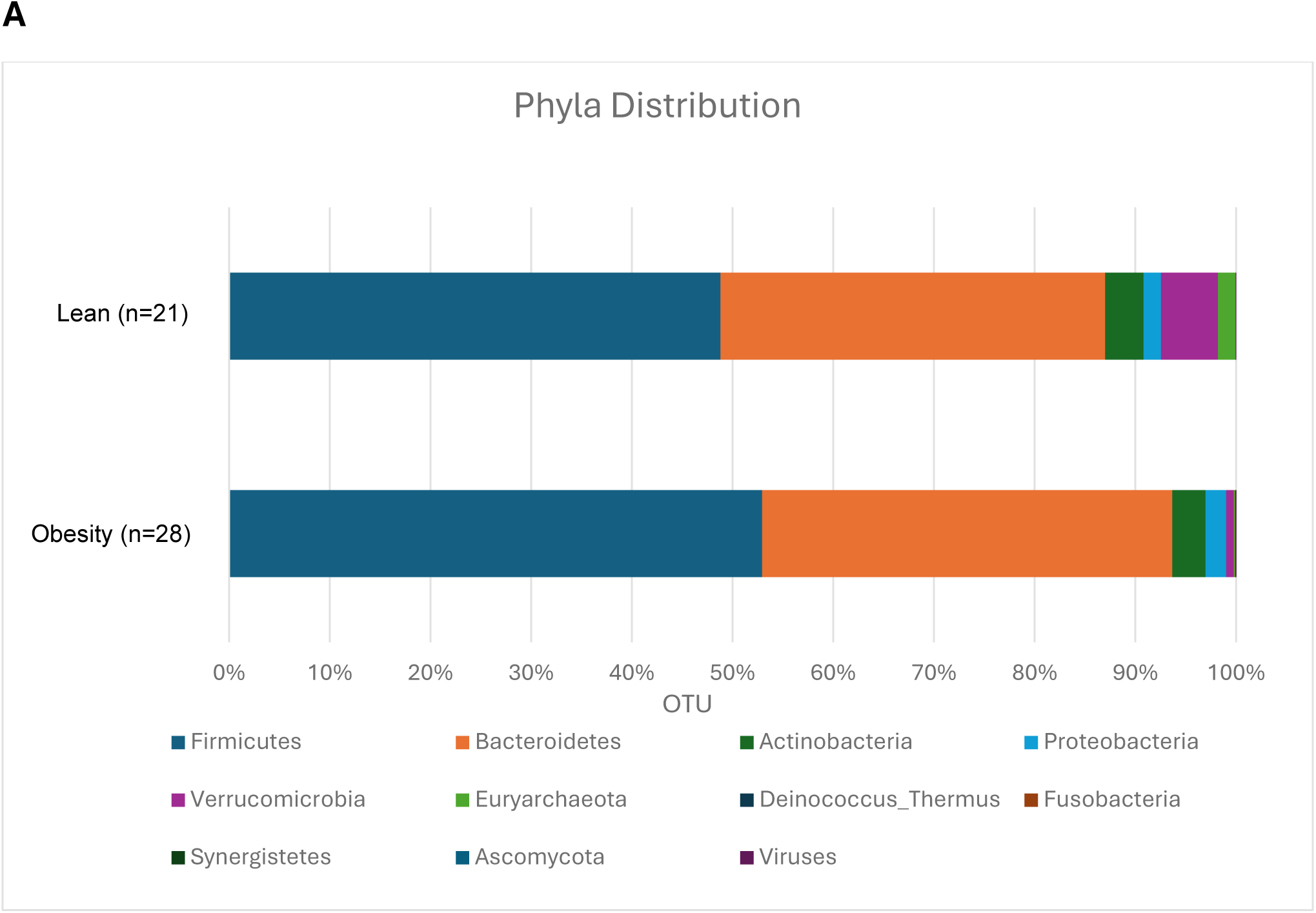
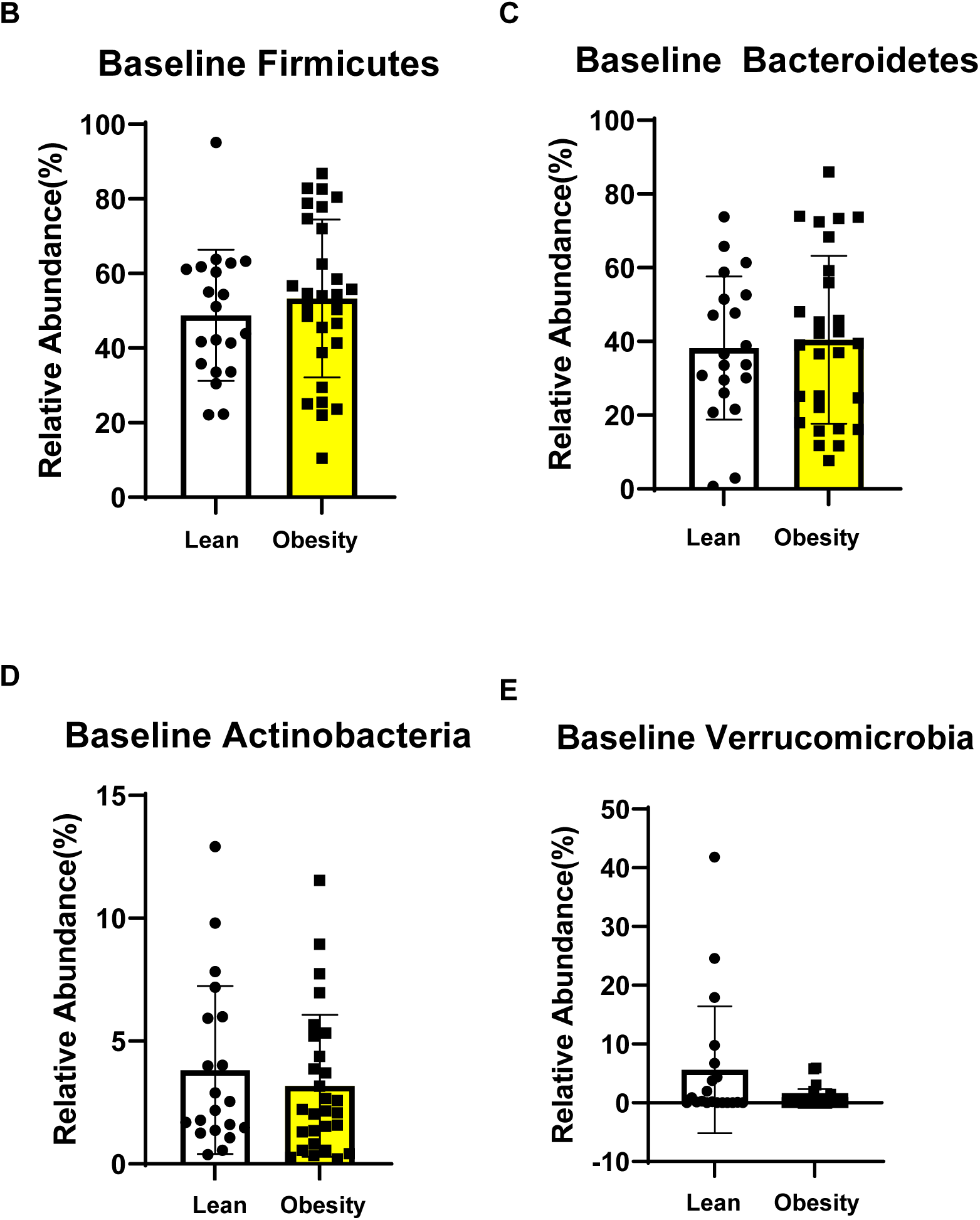
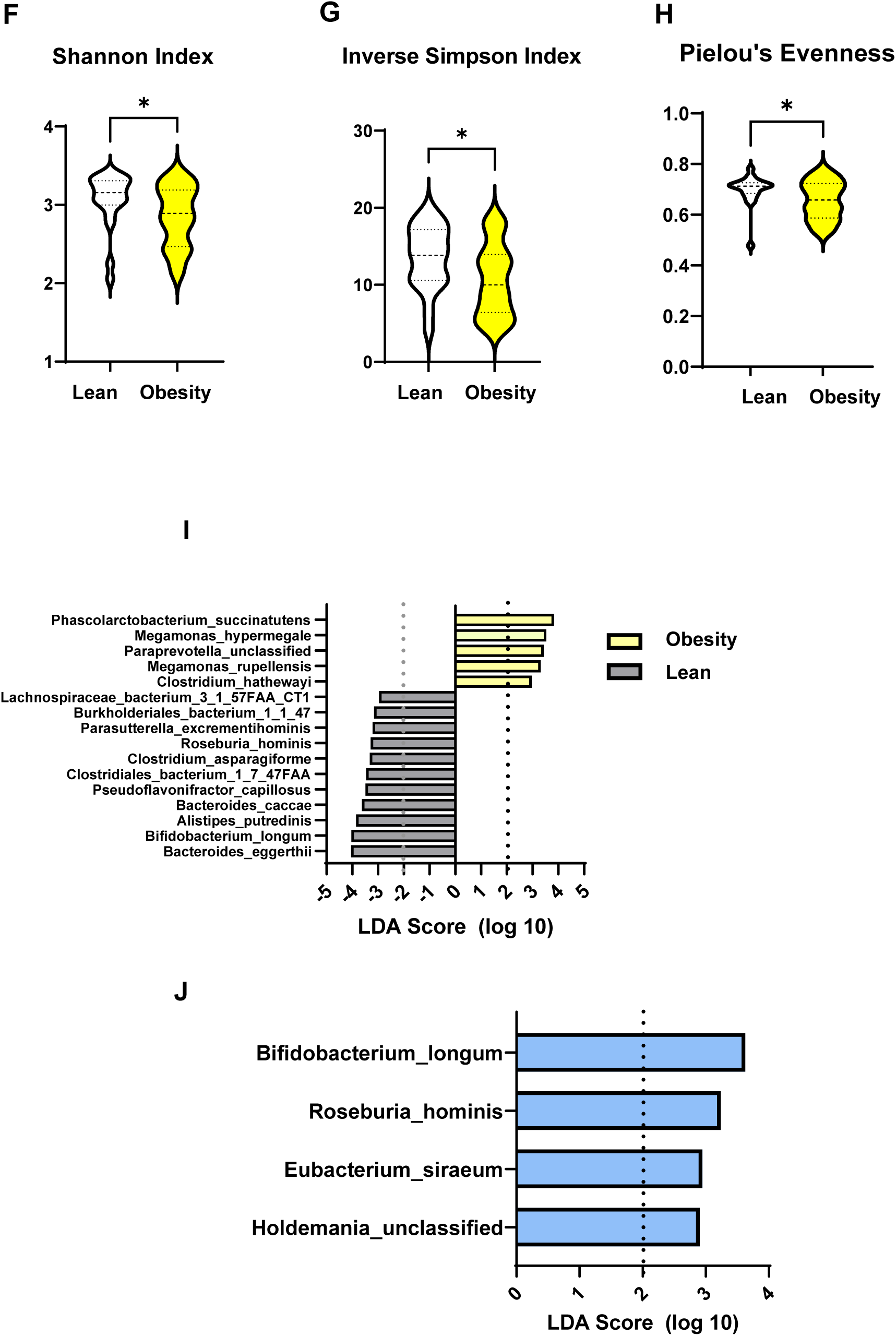
Baseline distribution of microbial phyla abundance in lean and obesity groups. (A-E) Comparison of enriched phyla in lean vs subjects with obesity, (F-H) measure of species diversity in lean vs subjects with obesity. Data are means ± SEM. *p<0.05. (I) Bacteria abundance in lean vs subjects with obesity at baseline. (J) Bacteria abundance after synbiotic treatment in obese subjects. Subjects with obesity are highlighted in yellow.

Paired (pre- and post-intervention) gut microbiome samples were available in 38 subjects. In lean subjects, none of the interventions resulted in significant changes in enrichment of *Firmicutes*, *Bacteroidetes*, *Actinobacteria*, or *Veruccomicrobia* (data not shown). In the obese group, the synbiotic caused significant enrichment of *Bifidobacterium longum*, *Roseburia hominis*, *Eubacterium siraeum*, and *Holdemania* genus bacteria (Figure 4J). Sevelamer did not cause significant enrichment of any bacteria species (not shown).

### Plasma metabolomics

Non-targeted plasma metabolomics was conducted to identify changes in metabolites that may help explain the improvement in insulin sensitivity and lipid levels seen in subjects with obesity that received sevelamer. Concentrations of 95 plasma metabolites were significantly (*q*<0.10) different between lean and obese groups at baseline; 85 metabolites were higher (Table S3A) and 10 were lower in the obese group (Table S3B). The heatmap shown in Figure 5A illustrates the differential abundance of various metabolites between lean and obese groups across multiple metabolic pathways. Prominent metabolomic differences in obese versus lean subjects include increased branched amino acids, fatty acids, acylcarnitines, diacylglycerols, ceramides, phospholipids, sphingomyelins, eicosanoids, endocannabinoids, and xenobiotics. Subjects in the obese group had an upregulation of pathways related to methionine, cysteine, S-Adenosyl-L-methionine (SAM), and taurine metabolism (cystathionine and cysteine sulfinic acid), as well as leucine, isoleucine, and valine metabolism (isovalerate).

**Figure 5.**
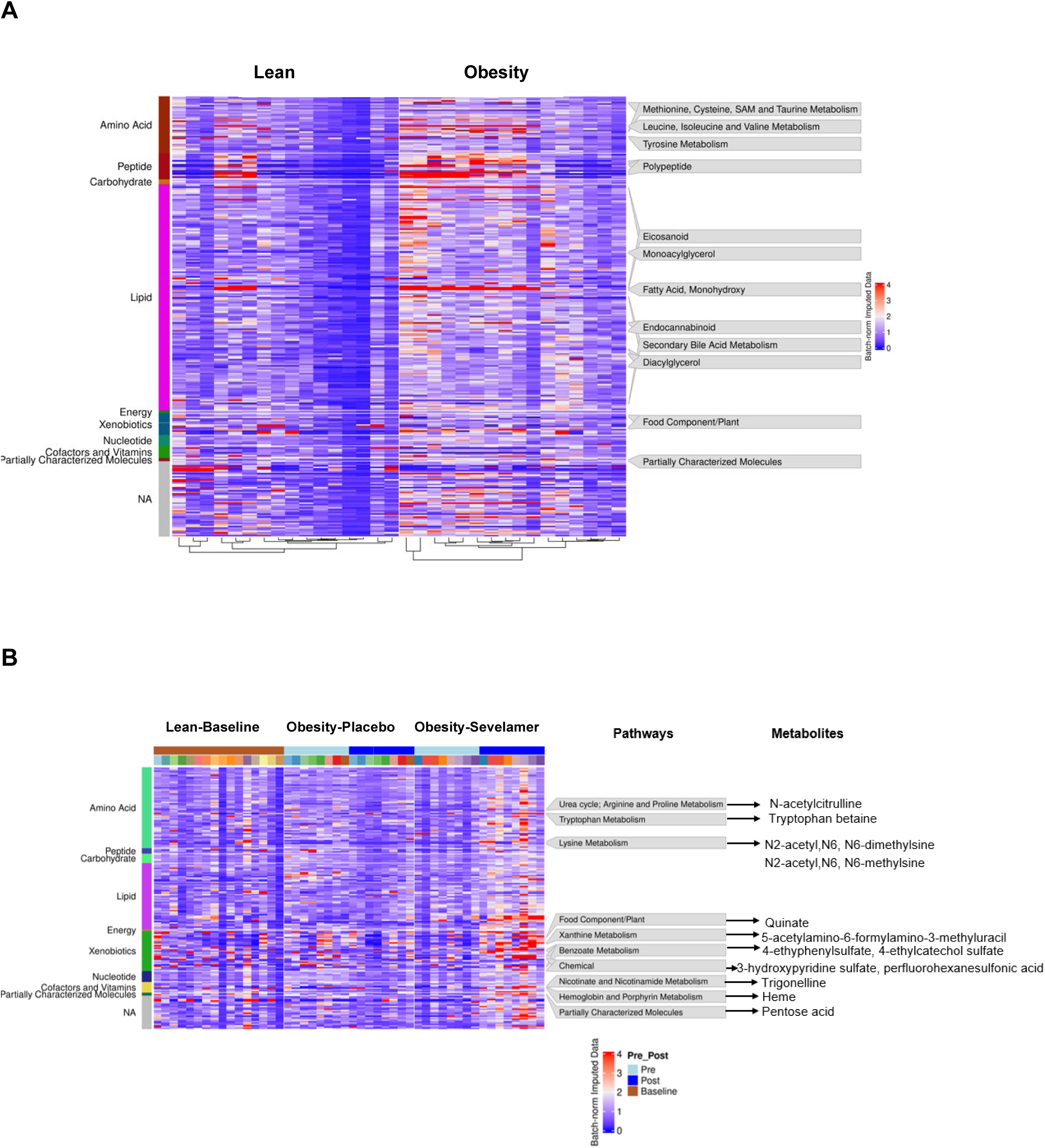
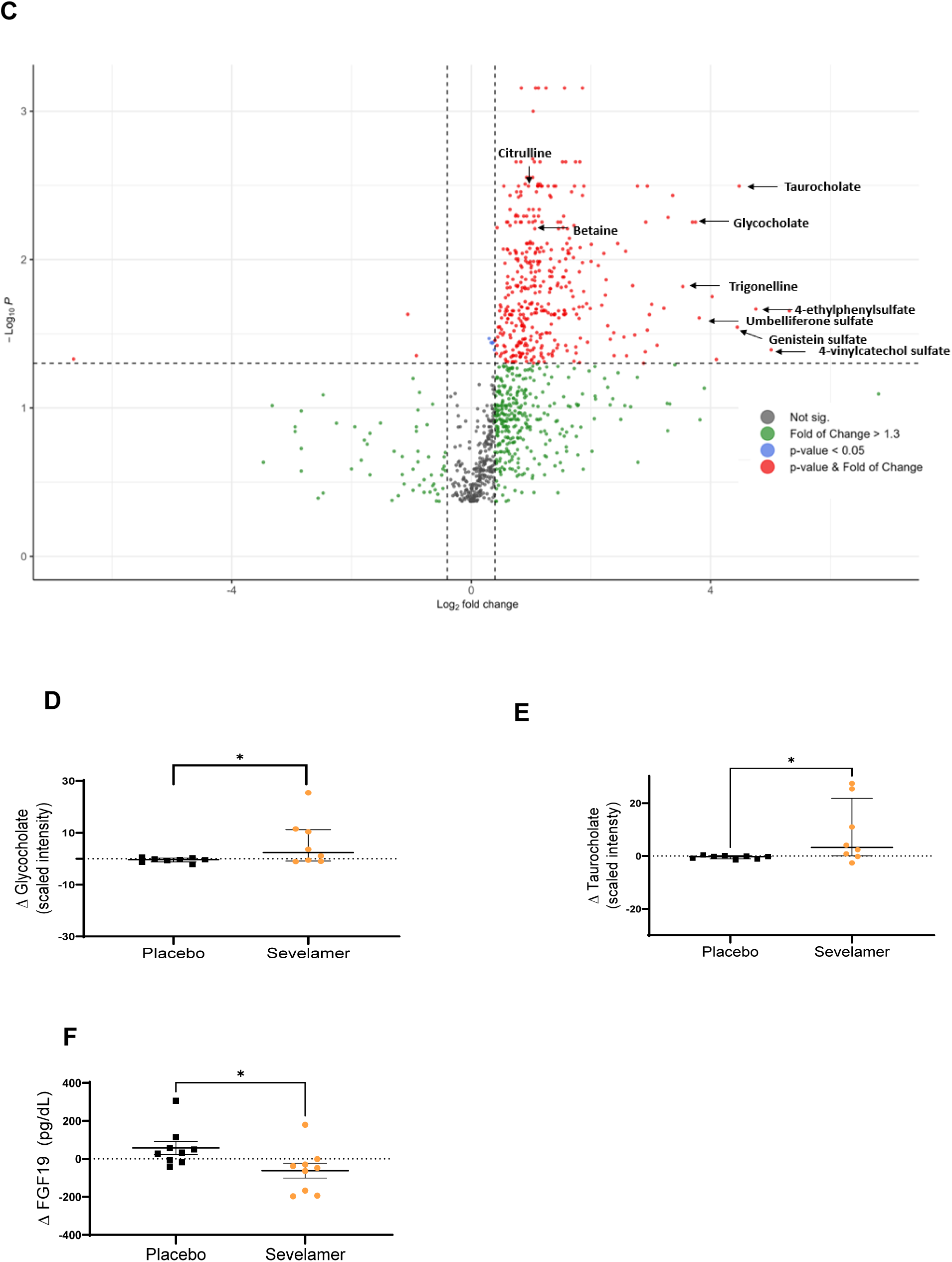
Heatmap and volcano plot of changes in metabolites and effect of placebo and sevelamer on plasma taurocholate, glycocholate and FGF19 concentration in the obesity group. (A) Heatmap of plasma metabolite levels in lean and obesity subjects. This heatmap visualizes the plasma concentrations of 286 significantly different metabolites between lean and obesity groups at baseline (Welch’s t-Test, p < 0.05). Three groups are included and labeled on the top bar: Lean at baseline (left section) and Obese before treatment (left section). Each column represents an individual participant. The rows represent individual metabolites, with examples of the top 10% most highly significant on the right side. The color intensity represents the normalized and imputed data for each metabolite, with the following gradient: Blue: Lower levels (value = 0); Red: Higher levels (value = 4); Intermediate colors (purple shades): Moderate levels (value = 2). The leftmost color bars represent different nutrients groupings for all the metabolites. (B) Heatmap of plasma metabolite levels in the Obesity group pre- and post-sevelamer. The heatmap visualizes plasma concentrations of 194 significantly different metabolites in the Sevelamer group during pre- vs. post-intervention analyses within the Obesity group (paired t-test, p < 0.05). Three groups are included and labeled on the top bar: Lean at baseline (left section, brown), Obesity receiving placebo [middle section, Pre (light blue) and Post (blue) group], and Obesity receiving sevelamer [right section, Pre (light blue) and Post (blue) group]. Each column represents an individual participant, with each color representing a different subject, and they are ranked in the same order in the Pre and Post groups. The rightmost rows represent individual metabolites, with examples of top 10% most highly significant listed on the right side along with their related pathway. The color intensity represents the normalized and imputed data for each metabolite, with the following gradient: Blue: Lower levels (value = 0); Red: Higher levels (value = 4); Intermediate colors (purple shades): Moderate levels (value = 2). The leftmost color bars represent different groupings for all the metabolites. (C) Volcano plot of changes in metabolite concentrations with sevelamer treatment in the obese group, (D) glycocholate, (E) taurocholate, and (F) FGF19 concentrations. Data are means ± SEM. *p<0.05.

Pre- vs. post-intervention paired comparisons in subjects with obesity revealed significant changes in 102 metabolites in the placebo group (Table S4) and 117 metabolites in the sevelamer group (Table S5). Comparisons between treatment groups revealed that 82 metabolites were higher and 5 were lower post-sevelamer vs. post-placebo (Tables S6A and S6B respectively). The top 10 percent of metabolites with most significant changes caused by sevelamer included metabolites involved in the urea cycle (N-acetylcitrulline) and metabolism of tryptophan (tryptophan betaine), lysine (N2-acetyl,N6,N6-dimethyllysine, N2-acetyl,N6-methyllysine), and nicotinamide [trigonelline (N’-methylnicotinate)]. Sevelamer treatment also increased the level of several food-related metabolites involved in metabolism of xanthine (5-acetylamino-6-formylamino-3-methyluracil) and benzoate [4-ethylphenylsulfate and 4-ethylcatechol sulfate] (Figure 5B). Sevelamer robustly increased the levels of other xenobiotics derived from plants including the isoflavone genistein (21-fold) and the coumarin derivative umbelliferone (14-fold) (Figures 5B and Fig. 5C). The concentrations of the gut-derived phenolics 3-methoxycatechol sulfate (13-fold) and 1,2,3-benzenetriol sulfate (39-fold) as well as 4-vinylcatechol sulfate (32-fold), highly abundant in coffee, and 4-ethylphenylsulfate (27-fold), which is produced by the gut microbiome, were all robustly increased by sevelamer.

Plasma taurocholate (Figure 5E) and glycocholate (Figure 5D) increased in the majority of subjects with obesity that received sevelamer. In view of changes in these bile acids, we measured FGF-19 concentrations because this protein is modulated by bile acid levels in the gut lumen. We found that sevelamer decreased FGF-19 concentrations (Figure 5F).

## DISCUSSION

Sevelamer is a non-absorbable, cross-linked polymer with high affinity for phosphates that is FDA-approved for use as a phosphate binder in patients with chronic kidney disease on dialysis. In a previous study, Bronden et al reported improvements in glycemia and LDL cholesterol in subjects with type 2 diabetes after a short (seven day) sevelamer course (16), although insulin sensitivity was not evaluated. In another study from the same group, they report a small reduction in HOMA-IR (17) in subjects with type 2 diabetes given sevelamer, effect that we did not observe in the present study. This discrepancy may be due to differences in study population. Moreover, we find in this study that sevelamer administration leads to a significant improvement in peripheral insulin sensitivity in non-diabetic subjects with obesity, as measured using insulin clamping, the gold standard for quantitating insulin sensitivity.

As expected, we found that subjects with obesity and insulin resistance had increased plasma LPS concentration. However, sevelamer did not affect LPS levels in this population of subjects with obesity. Patients with advanced kidney disease have elevated peripheral LPS concentrations and, both, cross-sectional (18) and prospective (19, 20) studies have shown that sevelamer reduces systemic LPS and markers of inflammation in this population. In animal models of obesity and metabolic dysfunction-associated steatotic liver disease (MASLD), sevelamer also reduces LPS concentrations and improves metabolic outcomes and inflammation (21). In contrast, in patients with HIV infection, who also have chronic endotoxemia, sevelamer failed to reduce LPS (22). The inconsistency in LPS response to sevelamer suggests that the ability of this drug to lower systemic LPS depends upon the underlying pathology.

Improvements in insulin resistance caused by sevelamer were not accompanied by changes in parameters of metabolic endotoxemia, intestinal permeability, or systemic inflammation, indicating that sevelamer works through an LPS-independent mechanism. Thus, we pursued a non-targeted metabolomics approach to help determine the mechanism by which sevelamer improves glucose and lipid metabolism. Plasma metabolomics revealed baseline differences between lean and obesity groups in numerous metabolites previously associated with insulin resistance, including branched amino acids, fatty acids, acylcarnitines, diacylglycerols, ceramides, phospholipids, sphingomyelins, eicosanoids, and endocannabinoids (23–34). Nonetheless, sevelamer did not affect levels of these metabolites, indicating that this drug does not improve insulin sensitivity and lipid levels by reverting concentrations of these metabolites in subjects with obesity to those in lean subjects.

Sevelamer increased plasma levels of taurocholic acid and glycocholic acid. In addition to binding free phosphate and LPS, sevelamer can bind to bile acids in the gut (35). In line with the study from Bronden et al (16), sevelamer reduced plasma fibroblast growth factor-19 (FGF-19) concentrations. FGF-19 production in the gut depends upon bile acid stimulation of intestinal farnesyl X receptor (FXR); thus, a reduction in plasma FGF-19 is indicative of lower free bile acid levels in the gut lumen. Because FGF-19 acts on the fibroblast growth factor receptor (FGFR)4 to decrease hepatic bile acid synthesis, a decrease in its levels would result in an increase in bile acid production and secretion, which is what we and others (16) observed. Inhibition of intestinal FXR has been shown to have beneficial impacts on insulin sensitivity in animal models of obesity (36). Thus, bile acid sequestration in the gut may contribute to the insulin-sensitizing and cholesterol-lowering effect of sevelamer. Yet, this mechanism is unlikely to solely explain improvements in peripheral glucose disposal since other bile acid sequestrants such as colesevelam do not improve insulin resistance (37).

In addition to bile acids, sevelamer administration to subjects with obesity led to an increase in the concentrations of several metabolites previously associated with positive changes in glucose and lipid metabolism. For example, sevelamer increased levels of citrulline, an amino acid found in fruits and vegetables and produced in the body as an intermediate within the urea cycle and as a by-product of arginine metabolism by nitric oxide synthase. Oral administration of citrulline increases nitric oxide production (38) and reduces blood glucose concentrations in patients with type 2 diabetes (39). Sevelamer also increased levels of betaine, which is found in vegetables and also produced by the body through oxidation of choline. Betaine administration improves insulin sensitivity in rodents (40) and glucose tolerance in humans (41). In addition, sevelamer increased levels of several other plant-derived compounds including quinate, trigonelline, genistein, and umbelliferone. Quinate activates AMP-activated protein kinase (AMPK) and decreases lipid levels (42) and trigonelline functions as an NAD+ precursor that improves mitochondrial function in muscle (43). Genistein is an isoflavone and phytoestrogen that also activates AMPK and improves glucose and lipid levels (44). Umbelliferone has both anti-inflammatory and glucose lowering properties (45).

Analysis of stool samples revealed baseline differences in the gut microbiome between the lean and obese groups. The gut microbiome of lean subjects is more diverse than subjects with obesity and shows preferential enrichment of health-promoting bacteria, including *Bifidobacterium longum.* Treatment with the synbiotic resulted in increased enrichment of *Bifidobacterium longum* in subjects with obesity. Some other bacteria including *Roseburia, Eurbacterium,* and *Holdemania* genus bacteria were also enriched after treatment with the synbiotic. Nonetheless, the synbiotic did not affect any metabolic or intestinal barrier parameters, indicating that enrichment of these bacterial species is not sufficient to improve metabolic outcomes in this population.

Supplementation for longer than one month may be necessary to improve metabolic parameters. Future studies may also evaluate the effect that enrichment and/or depletion of other bacterial species individually or in combination have on glucose metabolism.

In conclusion, sevelamer acts as an insulin-sensitizing and lipid-lowering drug, but the beneficial metabolic effects of this drug are not mediated by changes in the gut microbiome or metabolic endotoxemia. On the other hand, sevelamer caused changes in multiple metabolites that have been previously linked to improvements in glucose and lipid metabolism, including biliary acids, amino acids, and food compounds/xenobiotics. Identifying the specific compounds that mediate the metabolic effects of sevelamer could lead to the development of novel strategies to improve metabolic outcomes in insulin-resistant subjects.

## Data Availability

All data produced in the present study are available upon reasonable request to the authors.

## ACKNOWLEDGMENTS

*Bifidobacterium longum* and oligofructose were kindly provided by Lallemand, inc (Montreal, Canada). All de-identified data will be shared upon request to the corresponding author via email.

## Funding

This study was supported by grant 7-13-GSK-01 from the American Diabetes association and NIH grants R01DK080157, P30AG044271, P30AG013319, UL1TR002645, R01AG069690, and R01AG05269. Dr. Eric Baeuerle was funded in part by the South Texas MSTP (T32GM113896).

## Disclosure

The authors declared no conflicts of interest.

## Author contributions

N.M. conceived and planned the study. E.B., M.K.S., N.Z., S.E., N.M., and H.L. performed the experiments. E.B., M.K.S., J.N., N.Z., H.L., V.G., CP.W., and Z.Y. performed the analyses. E.B., M.K.S., J.N., N.Z. H.L., N.S., R.F., CP.W., Q.D., Z.Y., A.K., S.E. and N.M. interpreted data. E.B., M.K.S., and N.M. wrote the manuscript. All authors critically reviewed the manuscript for intellectual content and approved it for publication. N.M. acquired funding and is the guarantor of this work and, as such, had full access to all the data in the study and takes responsibility for the integrity of the data and the accuracy of the data analysis.

## Notes

### Competing Interest Statement

The authors have declared no competing interest.

### Clinical Trial

NCT02127125

### Funding Statement

American Diabetes Association
National Institutes of Health

### Author Declarations

Institutional Review Board of the University of Texas Health Science Center at San Antonio

## REFERENCES

1. Ley RE, Turnbaugh PJ, Klein S, Gordon JI. Microbial ecology: human gut microbes associated with obesity. Nature. 2006;444(7122):1022–3. Epub 2006/12/22. doi: 10.1038/4441022a. PubMed PMID: 17183309.

2. Turnbaugh PJ, Ridaura VK, Faith JJ, Rey FE, Knight R, Gordon JI. The effect of diet on the human gut microbiome: a metagenomic analysis in humanized gnotobiotic mice. Sci Transl Med. 2009;1(6):6ra14. Epub 2010/04/07. doi: 10.1126/scitranslmed.3000322. PubMed PMID: 20368178; PubMed Central PMCID: PMCPMC2894525.

3. Dabke K, Hendrick G, Devkota S. The gut microbiome and metabolic syndrome. The Journal of clinical investigation. 2019;129(10):4050–7.

4. Cani PD, Amar J, Iglesias MA, Poggi M, Knauf C, Bastelica D, et al. Metabolic endotoxemia initiates obesity and insulin resistance. Diabetes. 2007;56(7):1761–72. Epub 2007/04/26. doi: 10.2337/db06-1491. PubMed PMID: 17456850.

5. Liang H, Hussey SE, Sanchez-Avila A, Tantiwong P, Musi N. Effect of lipopolysaccharide on inflammation and insulin action in human muscle. PLoS One. 2013;8(5):e63983. Epub 2013/05/25. doi: 10.1371/journal.pone.0063983. PubMed PMID: 23704966; PubMed Central PMCID: PMCPMC3660322.

6. Pussinen PJ, Havulinna AS, Lehto M, Sundvall J, Salomaa V. Endotoxemia is associated with an increased risk of incident diabetes. Diabetes Care. 2011;34(2):392–7. Epub 2011/01/29. doi: 10.2337/dc10-1676. PubMed PMID: 21270197; PubMed Central PMCID: PMCPMC3024355.

7. Erridge C, Attina T, Spickett CM, Webb DJ. A high-fat meal induces low-grade endotoxemia: evidence of a novel mechanism of postprandial inflammation. Am J Clin Nutr. 2007;86(5):1286–92. Epub 2007/11/10. doi: 10.1093/ajcn/86.5.1286. PubMed PMID: 17991637.

8. Cani PD, Possemiers S, Van de Wiele T, Guiot Y, Everard A, Rottier O, et al. Changes in gut microbiota control inflammation in obese mice through a mechanism involving GLP-2-driven improvement of gut permeability. Gut. 2009;58(8):1091–103. Epub 2009/02/26. doi: 10.1136/gut.2008.165886. PubMed PMID: 19240062; PubMed Central PMCID: PMCPMC2702831.

9. Membrez M, Blancher F, Jaquet M, Bibiloni R, Cani PD, Burcelin RG, et al. Gut microbiota modulation with norfloxacin and ampicillin enhances glucose tolerance in mice. FASEB J. 2008;22(7):2416–26. Epub 2008/03/11. doi: 10.1096/fj.07-102723. PubMed PMID: 18326786.

10. Malaguarnera M, Vacante M, Antic T, Giordano M, Chisari G, Acquaviva R, et al. Bifidobacterium longum with fructo-oligosaccharides in patients with non alcoholic steatohepatitis. Dig Dis Sci. 2012;57(2):545–53. Epub 2011/09/09. doi: 10.1007/s10620-011-1887-4. PubMed PMID: 21901256.

11. Juby LD, Rothwell J, Axon AT. Lactulose/mannitol test: an ideal screen for celiac disease. Gastroenterology. 1989;96(1):79–85. Epub 1989/01/01. doi: 10.1016/0016-5085(89)90767-1. PubMed PMID: 2491824.

12. DeFronzo RA, Tobin JD, Andres R. Glucose clamp technique: a method for quantifying insulin secretion and resistance. Am J Physiol. 1979;237(3):E214–23. Epub 1979/09/01. doi: 10.1152/ajpendo.1979.237.3.E214. PubMed PMID: 382871.

13. Meyer M, Kircher M. Illumina sequencing library preparation for highly multiplexed target capture and sequencing. Cold Spring Harb Protoc. 2010;2010(6):pdb prot5448. Epub 2010/06/03. doi: 10.1101/pdb.prot5448. PubMed PMID: 20516186.

14. Oksanen J, Blanchet, F.G., Friendly, M., Kindt, R., Legendre, P., McGlinn, D., Minchin, P.R., O’Hara, R.B., Simpson, G.L., Solymos, P., Stevens, M.H.H., Szoecs, E., & Wagner, H. vegan: Community Ecology Package. R package version 2.5-6. 2.5-6 ed: R Foundation for Statistical Computing; 2019.

15. Segata N, Izard J, Waldron L, Gevers D, Miropolsky L, Garrett WS, et al. Metagenomic biomarker discovery and explanation. Genome Biol. 2011;12(6):R60. Epub 2011/06/28. doi: 10.1186/gb-2011-12-6-r60. PubMed PMID: 21702898; PubMed Central PMCID: PMCPMC3218848.

16. Bronden A, Mikkelsen K, Sonne DP, Hansen M, Vaben C, Gabe MN, et al. Glucose-lowering effects and mechanisms of the bile acid-sequestering resin sevelamer. Diabetes Obes Metab. 2018;20(7):1623–31. Epub 2018/03/02. doi: 10.1111/dom.13272. PubMed PMID: 29493868.

17. Nerild HH, Bronden A, Haddouchi AE, Ellegaard AM, Hartmann B, Rehfeld JF, et al. Elucidating the glucose-lowering effect of the bile acid sequestrant sevelamer. Diabetes Obes Metab. 2024;26(4):1252–63. Epub 2023/12/28. doi: 10.1111/dom.15421. PubMed PMID: 38151760.

18. Sun PP, Perianayagam MC, Jaber BL. Sevelamer hydrochloride use and circulating endotoxin in hemodialysis patients: a pilot cross-sectional study. J Ren Nutr. 2009;19(5):432–8. Epub 2009/05/26. doi: 10.1053/j.jrn.2009.01.022. PubMed PMID: 19464926.

19. Stinghen AE, Goncalves SM, Bucharles S, Branco FS, Gruber B, Hauser AB, et al. Sevelamer decreases systemic inflammation in parallel to a reduction in endotoxemia. Blood Purif. 2010;29(4):352–6. Epub 2010/04/02. doi: 10.1159/000302723. PubMed PMID: 20357435.

20. Navarro-Gonzalez JF, Mora-Fernandez C, Muros de Fuentes M, Donate-Correa J, Cazana-Perez V, Garcia-Perez J. Effect of phosphate binders on serum inflammatory profile, soluble CD14, and endotoxin levels in hemodialysis patients. Clin J Am Soc Nephrol. 2011;6(9):2272–9. Epub 2011/07/26. doi: 10.2215/CJN.01650211. PubMed PMID: 21784820; PubMed Central PMCID: PMCPMC3359006.

21. Tsuji Y, Kaji K, Kitade M, Kaya D, Kitagawa K, Ozutsumi T, et al. Bile Acid Sequestrant, Sevelamer Ameliorates Hepatic Fibrosis with Reduced Overload of Endogenous Lipopolysaccharide in Experimental Nonalcoholic Steatohepatitis. Microorganisms. 2020;8(6). Epub 2020/06/25. doi: 10.3390/microorganisms8060925. PubMed PMID: 32575352; PubMed Central PMCID: PMCPMC7357162.

22. Sandler NG, Zhang X, Bosch RJ, Funderburg NT, Choi AI, Robinson JK, et al. Sevelamer does not decrease lipopolysaccharide or soluble CD14 levels but decreases soluble tissue factor, low-density lipoprotein (LDL) cholesterol, and oxidized LDL cholesterol levels in individuals with untreated HIV infection. J Infect Dis. 2014;210(10):1549–54. Epub 2014/05/28. doi: 10.1093/infdis/jiu305. PubMed PMID: 24864123; PubMed Central PMCID: PMCPMC4215074.

23. Newgard CB, An J, Bain JR, Muehlbauer MJ, Stevens RD, Lien LF, et al. A branched-chain amino acid-related metabolic signature that differentiates obese and lean humans and contributes to insulin resistance. Cell Metab. 2009;9(4):311–26. Epub 2009/04/10. doi: 10.1016/j.cmet.2009.02.002. PubMed PMID: 19356713; PubMed Central PMCID: PMCPMC3640280.

24. Newgard CB. Interplay between lipids and branched-chain amino acids in development of insulin resistance. Cell Metab. 2012;15(5):606–14. Epub 2012/05/09. doi: 10.1016/j.cmet.2012.01.024. PubMed PMID: 22560213; PubMed Central PMCID: PMCPMC3695706.

25. Mihalik SJ, Goodpaster BH, Kelley DE, Chace DH, Vockley J, Toledo FG, et al. Increased levels of plasma acylcarnitines in obesity and type 2 diabetes and identification of a marker of glucolipotoxicity. Obesity (Silver Spring). 2010;18(9):1695–700. Epub 2010/01/30. doi: 10.1038/oby.2009.510. PubMed PMID: 20111019; PubMed Central PMCID: PMCPMC3984458.

26. Erion DM, Shulman GI. Diacylglycerol-mediated insulin resistance. Nat Med. 2010;16(4):400-2. Epub 2010/04/09. doi: 10.1038/nm0410-400. PubMed PMID: 20376053; PubMed Central PMCID: PMCPMC3730126.

27. Galadari S, Rahman A, Pallichankandy S, Galadari A, Thayyullathil F. Role of ceramide in diabetes mellitus: evidence and mechanisms. Lipids Health Dis. 2013;12:98. Epub 2013/07/10. doi: 10.1186/1476-511X-12-98. PubMed PMID: 23835113; PubMed Central PMCID: PMCPMC3716967.

28. Sokolowska E, Blachnio-Zabielska A. The Role of Ceramides in Insulin Resistance. Front Endocrinol (Lausanne). 2019;10:577. Epub 2019/09/10. doi: 10.3389/fendo.2019.00577. PubMed PMID: 31496996; PubMed Central PMCID: PMCPMC6712072.

29. Jornayvaz FR, Shulman GI. Diacylglycerol activation of protein kinase Cepsilon and hepatic insulin resistance. Cell Metab. 2012;15(5):574–84. Epub 2012/05/09. doi: 10.1016/j.cmet.2012.03.005. PubMed PMID: 22560210; PubMed Central PMCID: PMCPMC3353749.

30. Borkman M, Storlien LH, Pan DA, Jenkins AB, Chisholm DJ, Campbell LV. The relation between insulin sensitivity and the fatty-acid composition of skeletal-muscle phospholipids. N Engl J Med. 1993;328(4):238–44. Epub 1993/01/28. doi: 10.1056/NEJM199301283280404. PubMed PMID: 8418404.

31. Luo P, Wang MH. Eicosanoids, beta-cell function, and diabetes. Prostaglandins Other Lipid Mediat. 2011;95(1-4):1–10. Epub 2011/07/16. doi: 10.1016/j.prostaglandins.2011.06.001. PubMed PMID: 21757024; PubMed Central PMCID: PMCPMC3144311.

32. Straczkowski M, Kowalska I, Nikolajuk A, Dzienis-Straczkowska S, Kinalska I, Baranowski M, et al. Relationship between insulin sensitivity and sphingomyelin signaling pathway in human skeletal muscle. Diabetes. 2004;53(5):1215–21. Epub 2004/04/28. doi: 10.2337/diabetes.53.5.1215. PubMed PMID: 15111489.

33. Green CD, Maceyka M, Cowart LA, Spiegel S. Sphingolipids in metabolic disease: The good, the bad, and the unknown. Cell Metab. 2021;33(7):1293–306. Epub 2021/07/08. doi: 10.1016/j.cmet.2021.06.006. PubMed PMID: 34233172; PubMed Central PMCID: PMCPMC8269961.

34. Ge Q, Maury E, Rycken L, Gerard J, Noel L, Detry R, et al. Endocannabinoids regulate adipokine production and the immune balance of omental adipose tissue in human obesity. Int J Obes (Lond). 2013;37(6):874–80. Epub 2012/08/08. doi: 10.1038/ijo.2012.123. PubMed PMID: 22868830.

35. Braunlin W, Zhorov E, Guo A, Apruzzese W, Xu Q, Hook P, et al. Bile acid binding to sevelamer HCl. Kidney Int. 2002;62(2):611–9. Epub 2002/07/12. doi: 10.1046/j.1523-1755.2002.00459.x. PubMed PMID: 12110025.

36. Jiang C, Xie C, Lv Y, Li J, Krausz KW, Shi J, et al. Intestine-selective farnesoid X receptor inhibition improves obesity-related metabolic dysfunction. Nat Commun. 2015;6:10166. Epub 2015/12/17. doi: 10.1038/ncomms10166. PubMed PMID: 26670557; PubMed Central PMCID: PMCPMC4682112.

37. Henry RR, Aroda VR, Mudaliar S, Garvey WT, Chou HS, Jones MR. Effects of colesevelam on glucose absorption and hepatic/peripheral insulin sensitivity in patients with type 2 diabetes mellitus. Diabetes Obes Metab. 2012;14(1):40–6. Epub 2011/08/13. doi: 10.1111/j.1463-1326.2011.01486.x. PubMed PMID: 21831167; PubMed Central PMCID: PMCPMC4955577.

38. Theodorou AA, Zinelis PT, Malliou VJ, Chatzinikolaou PN, Margaritelis NV, Mandalidis D, et al. Acute L-Citrulline Supplementation Increases Nitric Oxide Bioavailability but Not Inspiratory Muscle Oxygenation and Respiratory Performance. Nutrients. 2021;13(10). Epub 2021/10/24. doi: 10.3390/nu13103311. PubMed PMID: 34684312; PubMed Central PMCID: PMCPMC8537281.

39. Abbaszadeh F, Azizi S, Mobasseri M, Ebrahimi-Mameghani M. The effects of citrulline supplementation on meta-inflammation and insulin sensitivity in type 2 diabetes: a randomized, double-blind, placebo-controlled trial. Diabetol Metab Syndr. 2021;13(1):52. Epub 2021/05/07. doi: 10.1186/s13098-021-00669-w. PubMed PMID: 33952324; PubMed Central PMCID: PMCPMC8097832.

40. Szkudelska K, Szkudelski T. The anti-diabetic potential of betaine. Mechanisms of action in rodent models of type 2 diabetes. Biomed Pharmacother. 2022;150:112946. Epub 2022/04/13. doi: 10.1016/j.biopha.2022.112946. PubMed PMID: 35413601.

41. Grizales AM, Patti ME, Lin AP, Beckman JA, Sahni VA, Cloutier E, et al. Metabolic Effects of Betaine: A Randomized Clinical Trial of Betaine Supplementation in Prediabetes. J Clin Endocrinol Metab. 2018;103(8):3038–49. Epub 2018/06/04. doi: 10.1210/jc.2018-00507. PubMed PMID: 29860335; PubMed Central PMCID: PMCPMC6692715.

42. Sung YY, Kim DS, Kim HK. Akebia quinata extract exerts anti-obesity and hypolipidemic effects in high-fat diet-fed mice and 3T3-L1 adipocytes. J Ethnopharmacol. 2015;168:17–24. Epub 2015/04/04. doi: 10.1016/j.jep.2015.03.051. PubMed PMID: 25835369.

43. Membrez M, Migliavacca E, Christen S, Yaku K, Trieu J, Lee AK, et al. Trigonelline is an NAD(+) precursor that improves muscle function during ageing and is reduced in human sarcopenia. Nat Metab. 2024;6(3):433–47. Epub 2024/03/20. doi: 10.1038/s42255-024-00997-x. PubMed PMID: 38504132; PubMed Central PMCID: PMCPMC10963276 M.J.S., S.M., V.S. and J.N.F. are or were employees of Nestle Research, which is part of the Societe des Produits Nestle S.A. L.G.K. was an employee of Nestle Health Sciences. K.M.G. has received reimbursement for speaking at conferences sponsored by companies selling nutritional products and is part of an academic consortium that has received research funding from BenevolentAI, Nestle Research and Danone. The other authors declare no competing interests.

44. Cederroth CR, Vinciguerra M, Gjinovci A, Kuhne F, Klein M, Cederroth M, et al. Dietary phytoestrogens activate AMP-activated protein kinase with improvement in lipid and glucose metabolism. Diabetes. 2008;57(5):1176–85. Epub 2008/04/19. doi: 10.2337/db07-0630. PubMed PMID: 18420492.

45. Yin J, Wang H, Lu G. Umbelliferone alleviates hepatic injury in diabetic db/db mice via inhibiting inflammatory response and activating Nrf2-mediated antioxidant. Biosci Rep. 2018;38(4). Epub 2018/07/04. doi: 10.1042/BSR20180444. PubMed PMID: 29967293; PubMed Central PMCID: PMCPMC6131207.

