## Supplementary Information for "Effect of Sevelamer versus *Bifidobacterium longum* on Insulin Sensitivity in Subjects with Obesity"

**Supplementary Materials**

Table S1. Subject characteristics. Data are means ± SD. *p<0.05

| **Parameters** | **Lean (N=22)**  Mean (SD) or N (%) | **Obesity (N=28)**  Mean (SD) or N (%) |
| --- | --- | --- |
| Age, years | 45.4 (3.0) | 51.9 ± 1.7 |
| Female, n (%), Sex (Male/Female) | 15 (68.2) | 20 (71.4) |
| Weight, kg | 65.2 (2.1) | 89.6 (2.0)* |
| Body mass index, kg/m^2^ | 23.0 (0.5) | 32.7 (0.4)* |
| Fasting plasma glucose, mmol/L | 5.22 (0.10) | 5.50 (0.07)* |
| 2 hrs glucose during OGTT, mmol/L | 5.68 (0.18) | 6.06 (0.19) |
| Hemoglobin A1c, % | 5.32 (0.07) | 5.49 (0.6) |

Table S2. Adverse events reported by subjects.

|  | Placebo | Sevelamer | Synbiotic |
| --- | --- | --- | --- |
| Abdominal discomfort | 3 | 3 | 3 |
| Lightheadedness | 2 | - | 1 |
| Nausea | 2 | - | - |
| Fatigue | - | 2 | - |
| Pruritis | 1 | - | - |
| Decreased appetite | - | 1 | - |
| Fibromyalgia | - | 1 | - |
| Shoulder pain | 1 | - | - |
| Groin discomfort | 1 | - | - |
| Constipation | 1 | - | - |
| Knee edema | - | 1 | - |
| Muscle contraction | - | - | 2 |
| Toothache | - | - | 1 |
| Headache | 1 | - | - |
| Abdominal itching | - | 1 | - |
| Palpitations | - | 1 | - |
| Ear discomfort | - | 1 | - |
| Allergic rhinitis | - | 1 | - |
| Total number of adverse events | 12 | 12 | 7 |

Table S3A. Baseline plasma metabolites higher in obesity vs. lean group (*q<0.1*)

| **Metabolite** | **Family** | **Sub-Pathway** | **Fold change** | **q-value** |
| --- | --- | --- | --- | --- |
| HWESASLLR | Peptide | Polypeptide | 7.32 | 0.0280 |
| erythritol | Xenobiotics | Food Component/Plant | 7.23 | 0.0650 |
| 5-hydroxyindole sulfate | Amino Acid | Tryptophan Metabolism | 6.62 | 0.0990 |
| bradykinin | Peptide | Polypeptide | 4.95 | 0.0391 |
| leukotriene B4 | Lipid | Eicosanoid | 4.09 | 0.0229 |
| 1-linolenoylglycerol (18:3) | Lipid | Monoacylglycerol | 3.89 | 0.0015 |
| 5-HETE | Lipid | Eicosanoid | 3.63 | 0.0207 |
| XHWESASXXR | Peptide | Polypeptide | 3.57 | 0.0675 |
| isoursodeoxycholate | Lipid | Secondary Bile Acid Metabolism | 3.46 | 0.0780 |
| 13-HODE + 9-HODE | Lipid | Fatty Acid, Monohydroxy | 3.30 | 0.0205 |
| cystathionine | Amino Acid | Methionine, Cysteine, SAM and Taurine Metabolism | 3.13 | 0.0500 |
| isovalerate (i5:0) | Amino Acid | Leucine, Isoleucine and Valine Metabolism | 3.11 | 0.0448 |
| metabolonic lactone sulfate | Partially Characterized Molecules | Partially Characterized Molecules | 3.06 | 0.0096 |
| palmitoyl-oleoyl-glycerol (16:0/18:1) [2] | Lipid | Diacylglycerol | 3.00 | 0.0149 |
| N-formylphenylalanine | Amino Acid | Tyrosine Metabolism | 2.94 | 0.0358 |
| HWESASXX | Peptide | Polypeptide | 2.89 | 0.0358 |
| HXGXA | Peptide | Polypeptide | 2.89 | 0.0813 |
| cysteine sulfinic acid | Amino Acid | Methionine, Cysteine, SAM and Taurine Metabolism | 2.87 | 0.0243 |
| N-linoleoyltaurine | Lipid | Endocannabinoid | 2.86 | 0.0485 |
| linoleoyl-linolenoyl-glycerol (18:2/18:3) [1] | Lipid | Diacylglycerol | 2.74 | 0.0084 |
| 5-HEPE | Lipid | Eicosanoid | 2.72 | 0.0275 |
| deoxycholic acid glucuronide | Lipid | Secondary Bile Acid Metabolism | 2.72 | 0.0378 |
| myristoyl-linoleoyl-glycerol (14:0/18:2) [2] | Lipid | Diacylglycerol | 2.70 | 0.0065 |
| trans-3,4-methyleneheptanoate | Xenobiotics | Food Component/Plant | 2.63 | 0.0210 |
| linoleoyl ethanolamide | Lipid | Endocannabinoid | 2.60 | 0.0243 |
| 1-linoleoylglycerol (18:2) | Lipid | Monoacylglycerol | 2.58 | 0.0122 |
| gamma-glutamylisoleucine | Peptide | Gamma-glutamyl Amino Acid | 2.58 | 0.0302 |
| palmitoyl-linoleoyl-glycerol (16:0/18:2) [2] | Lipid | Diacylglycerol | 2.52 | 0.0011 |
| 4-hydroxyglutamate | Amino Acid | Glutamate Metabolism | 2.46 | 0.0302 |
| leukotriene B5 | Lipid | Eicosanoid | 2.46 | 0.0586 |
| glutamate | Amino Acid | Glutamate Metabolism | 2.42 | 0.0096 |
| butyrate/isobutyrate (4:0) | Lipid | Short Chain Fatty Acid | 2.40 | 0.0449 |
| 2-linoleoylglycerol (18:2) | Lipid | Monoacylglycerol | 2.39 | 0.0220 |
| gamma-glutamylglutamate | Peptide | Gamma-glutamyl Amino Acid | 2.38 | 0.0205 |
| cis-3,4-methyleneheptanoylcarnitine | Lipid | Fatty Acid Metabolism (Acyl Carnitine, Medium Chain) | 2.37 | 0.0145 |
| palmitoyl-linoleoyl-glycerol (16:0/18:2) [1] | Lipid | Diacylglycerol | 2.36 | 0.0017 |
| diacylglycerol (16:1/18:2 [2], 16:0/18:3 [1]) | Lipid | Diacylglycerol | 2.32 | 0.0122 |
| 1-oleoylglycerol (18:1) | Lipid | Monoacylglycerol | 2.30 | 0.0158 |
| gamma-glutamylvaline | Peptide | Gamma-glutamyl Amino Acid | 2.30 | 0.0243 |
| 1-palmitoleoylglycerol (16:1) | Lipid | Monoacylglycerol | 2.28 | 0.0220 |
| 3-indoleglyoxylic acid | Xenobiotics | Food Component/Plant | 2.27 | 0.0275 |
| linoleoyl-linolenoyl-glycerol (18:2/18:3) [2] | Lipid | Diacylglycerol | 2.25 | 0.0084 |
| gamma-glutamylphenylalanine | Peptide | Gamma-glutamyl Amino Acid | 2.25 | 0.0280 |
| histidylalanine | Peptide | Dipeptide | 2.24 | 0.0676 |
| palmitoyl-oleoyl-glycerol (16:0/18:1) [1] | Lipid | Diacylglycerol | 2.23 | 0.0131 |
| retinal | Cofactors and Vitamins | Vitamin A Metabolism | 2.16 | 0.0220 |
| linoleoyl-linoleoyl-glycerol (18:2/18:2) [2] | Lipid | Diacylglycerol | 2.13 | 0.0122 |
| linoleoyl-linoleoyl-glycerol (18:2/18:2) [1] | Lipid | Diacylglycerol | 2.11 | 0.0380 |
| eicosapentaenoate (EPA; 20:5n3) | Lipid | Long Chain Polyunsaturated Fatty Acid (n3 and n6) | 2.11 | 0.0578 |
| 1-stearoyl-GPG (18:0) | Lipid | Lysophospholipid | 2.08 | 0.0586 |
| oleoyl-oleoyl-glycerol (18:1/18:1) [1] | Lipid | Diacylglycerol | 2.06 | 0.0280 |
| oleoyl-oleoyl-glycerol (18:1/18:1) [2] | Lipid | Diacylglycerol | 2.04 | 0.0280 |
| glycine conjugate of C10H14O2 (1) | Partially Characterized Molecules | Partially Characterized Molecules | 2.03 | 0.0229 |
| gamma-glutamyltyrosine | Peptide | Gamma-glutamyl Amino Acid | 2.02 | 0.0449 |
| gamma-glutamylthreonine | Peptide | Gamma-glutamyl Amino Acid | 1.99 | 0.0391 |
| pimelate (C7-DC) | Lipid | Fatty Acid, Dicarboxylate | 1.98 | 0.0264 |
| cis-3,4-methyleneheptanoylglycine | Lipid | Fatty Acid Metabolism (Acyl Glycine) | 1.98 | 0.0277 |
| sphingomyelin (d18:0/20:0, d16:0/22:0) | Lipid | Dihydrosphingomyelins | 1.97 | 0.0448 |
| glucuronide of piperine metabolite C17H21NO3 (4) | Xenobiotics | Food Component/Plant | 1.96 | 0.0378 |
| 1-stearoyl-GPI (18:0) | Lipid | Lysophospholipid | 1.96 | 0.0500 |
| bradykinin, des-arg(9) | Peptide | Polypeptide | 1.96 | 0.0676 |
| 1-palmitoylglycerol (16:0) | Lipid | Monoacylglycerol | 1.95 | 0.0500 |
| 5-methyluridine (ribothymidine) | Nucleotide | Pyrimidine Metabolism, Uracil containing | 1.94 | 0.0813 |
| N-acetyl-aspartyl-glutamate (NAAG) | Amino Acid | Glutamate Metabolism | 1.93 | 0.0870 |
| dihomo-linoleoylcarnitine (C20:2) | Lipid | Fatty Acid Metabolism (Acyl Carnitine, Polyunsaturated) | 1.91 | 0.0149 |
| 2-palmitoleoyl-GPC (16:1) | Lipid | Lysophospholipid | 1.91 | 0.0534 |
| oleoyl-linoleoyl-glycerol (18:1/18:2) [1] | Lipid | Diacylglycerol | 1.90 | 0.0065 |
| 1-stearoyl-GPC (18:0) | Lipid | Lysophospholipid | 1.90 | 0.0228 |
| 3-carboxy-4-methyl-5-pentyl-2-furanpropionate (3-CMPFP) | Lipid | Fatty Acid, Dicarboxylate | 1.90 | 0.0244 |
| oleoyl-linoleoyl-glycerol (18:1/18:2) [2] | Lipid | Diacylglycerol | 1.89 | 0.0084 |
| 1-(1-enyl-oleoyl)-GPE (P-18:1) | Lipid | Lysoplasmalogen | 1.89 | 0.0325 |
| arachidonate (20:4n6) | Lipid | Long Chain Polyunsaturated Fatty Acid (n3 and n6) | 1.89 | 0.0358 |
| N-palmitoyl-sphinganine (d18:0/16:0) | Lipid | Dihydroceramides | 1.89 | 0.0534 |
| docosahexaenoate (DHA; 22:6n3) | Lipid | Long Chain Polyunsaturated Fatty Acid (n3 and n6) | 1.88 | 0.0448 |
| 1-palmitoyl-GPI (16:0) | Lipid | Lysophospholipid | 1.87 | 0.0448 |
| N1-methylinosine | Nucleotide | Purine Metabolism, (Hypo)Xanthine/Inosine containing | 1.85 | 0.0275 |
| 1-palmitoleoyl-GPC (16:1)* | Lipid | Lysophospholipid | 1.85 | 0.0358 |
| caproate (6:0) | Lipid | Medium Chain Fatty Acid | 1.84 | 0.0576 |
| 1-linoleoyl-GPG (18:2)* | Lipid | Lysophospholipid | 1.82 | 0.0220 |
| sphingomyelin (d18:2/14:0, d18:1/14:1)* | Lipid | Sphingomyelins | 1.82 | 0.0392 |
| tetradecanedioate (C14-DC) | Lipid | Fatty Acid, Dicarboxylate | 1.82 | 0.0448 |
| gamma-glutamylleucine | Peptide | Gamma-glutamyl Amino Acid | 1.82 | 0.0477 |
| N-oleoyltaurine | Lipid | Endocannabinoid | 1.81 | 0.0358 |
| ceramide (d16:1/24:1, d18:1/22:1)* | Lipid | Ceramides | 1.81 | 0.0448 |
| methionine sulfoxide | Amino Acid | Methionine, Cysteine, SAM and Taurine Metabolism | 1.81 | 0.0576 |

Table S3B. Baseline plasma metabolites lower in obesity vs. lean group (*q<0.1*)

| **Metabolite** | **Family** | **Sub-Pathway** | **Fold change** | **q-value** |
| --- | --- | --- | --- | --- |
| heme | Cofactors and Vitamins | Hemoglobin and Porphyrin Metabolism | 0.15 | 0.0559 |
| 2-methoxyresorcinol sulfate | Xenobiotics | Chemical | 0.20 | 0.0951 |
| 4-methylguaiacol sulfate | Xenobiotics | Benzoate Metabolism | 0.21 | 0.0578 |
| p-cresol glucuronide* | Amino Acid | Tyrosine Metabolism | 0.23 | 0.0118 |
| glyco-beta-muricholate | Lipid | Primary Bile Acid Metabolism | 0.28 | 0.0500 |
| 4-methylcatechol sulfate | Xenobiotics | Benzoate Metabolism | 0.36 | 0.0557 |
| taurolithocholate 3-sulfate | Lipid | Secondary Bile Acid Metabolism | 0.36 | 0.0830 |
| p-cresol sulfate | Xenobiotics | Benzoate Metabolism | 0.48 | 0.0399 |
| hypotaurine | Amino Acid | Methionine, Cysteine, SAM and Taurine Metabolism | 0.51 | 0.0586 |
| ascorbic acid 3-sulfate | Cofactors and Vitamins | Ascorbate and Aldarate Metabolism | 0.55 | 0.0734 |

Table S4A. Plasma metabolites increased in obesity group pre- vs. post-placebo (*q<0.1*)

| **Metabolite** | **Family** | **Sub-Pathway** | **Fold change** | **q-value** |
| --- | --- | --- | --- | --- |
| hydroxybupropion | Xenobiotics | Drug - Psychoactive | 104.21 | 0.0869 |
| bilirubin (Z,Z) | Cofactors and Vitamins | Hemoglobin and Porphyrin Metabolism | 32.45 | 0.0024 |
| bilirubin degradation product, C17H18N2O4 (2) | Partially Characterized Molecules | Partially Characterized Molecules | 19.70 | 0.0002 |
| bilirubin degradation product, C17H18N2O4 (1) | Partially Characterized Molecules | Partially Characterized Molecules | 15.75 | 0.0016 |
| bilirubin degradation product, C17H18N2O4 (3) | Partially Characterized Molecules | Partially Characterized Molecules | 15.47 | 0.0001 |
| beta-cryptoxanthin | Cofactors and Vitamins | Vitamin A Metabolism | 9.51 | 0.0180 |
| heme | Cofactors and Vitamins | Hemoglobin and Porphyrin Metabolism | 8.94 | 0.0030 |
| pregnanediol-3-glucuronide | Lipid | Progestin Steroids | 8.01 | 0.0781 |
| alpha-tocopherol | Cofactors and Vitamins | Tocopherol Metabolism | 6.38 | 0.0531 |
| 3-hydroxystachydrine | Xenobiotics | Food Component/Plant | 5.53 | 0.0239 |
| bilirubin (E,E) | Cofactors and Vitamins | Hemoglobin and Porphyrin Metabolism | 5.24 | 0.0060 |
| alpha-ketobutyrate | Amino Acid | Methionine, Cysteine, SAM and Taurine Metabolism | 5.09 | 0.0022 |
| carotene diol (1) | Cofactors and Vitamins | Vitamin A Metabolism | 5.07 | 0.0350 |
| carotene diol (3) | Cofactors and Vitamins | Vitamin A Metabolism | 4.85 | 0.0463 |

Table S4B. Plasma metabolites decreased in obesity group pre- vs. post-placebo (*q<0.1*)

| **Metabolite** | **Family** | **Sub-Pathway** | **Fold change** | **q-value** |
| --- | --- | --- | --- | --- |
| Fibrinopeptide A* | Peptide | Fibrinogen Cleavage Peptide | 4.5 x 10^-3^ | 0.0001 |
| HXGXA* | Peptide | Polypeptide | 3.8 x 10^-3^ | 0.0002 |
| Fibrinopeptide A, phosphono-ser(3)* | Peptide | Fibrinogen Cleavage Peptide | 0.01 | 0.0002 |
| Fibrinopeptide B | Peptide | Fibrinogen Cleavage Peptide | 0.01 | 0.0002 |
| Fibrinopeptide B (1-13)** | Peptide | Fibrinogen Cleavage Peptide | 0.01 | 0.0002 |
| Fibrinopeptide A, des-ala(1)* | Peptide | Fibrinogen Cleavage Peptide | 0.01 | 0.0003 |
| leucylglycine | Peptide | Dipeptide | 0.01 | 0.0007 |
| histidylalanine | Peptide | Dipeptide | 0.02 | 0.0021 |
| bradykinin, des-arg(9) | Peptide | Polypeptide | 0.03 | 0.0003 |
| valylleucine | Peptide | Dipeptide | 0.03 | 0.0154 |
| dimethyl sulfone | Xenobiotics | Chemical | 0.04 | 0.0782 |
| XHWESASXXR* | Peptide | Polypeptide | 0.07 | 0.0398 |
| glycoursodeoxycholic acid sulfate (1) | Lipid | Secondary Bile Acid Metabolism | 0.08 | 0.0960 |
| 4-guanidinobutanoate | Amino Acid | Guanidino and Acetamido Metabolism | 0.09 | 0.0009 |
| glutamine conjugate of C6H10O2 (2)* | Partially Characterized Molecules | Partially Characterized Molecules | 0.09 | 0.0019 |
| bradykinin | Peptide | Polypeptide | 0.09 | 0.0210 |
| valylglycine | Peptide | Dipeptide | 0.10 | 0.0003 |
| HWESASXX* | Peptide | Polypeptide | 0.10 | 0.0009 |
| 5-HETE | Lipid | Eicosanoid | 0.10 | 0.0010 |
| isovalerate (i5:0) | Amino Acid | Leucine, Isoleucine and Valine Metabolism | 0.11 | 0.0025 |
| 13-HODE + 9-HODE | Lipid | Fatty Acid, Monohydroxy | 0.12 | 0.0013 |
| leukotriene B4 | Lipid | Eicosanoid | 0.13 | 0.0030 |
| 5-HEPE | Lipid | Eicosanoid | 0.16 | 0.0028 |
| 5alpha-androstan-3alpha,17beta-diol monosulfate (2) | Lipid | Androgenic Steroids | 0.17 | 0.0649 |
| leukotriene B5 | Lipid | Eicosanoid | 0.19 | 0.0019 |
| HWESASLLR | Peptide | Polypeptide | 0.19 | 0.0350 |
| gamma-glutamylisoleucine* | Peptide | Gamma-glutamyl Amino Acid | 0.20 | 0.0008 |
| glutamine conjugate of C7H12O2* | Partially Characterized Molecules | Partially Characterized Molecules | 0.20 | 0.0260 |
| butyrate/isobutyrate (4:0) | Lipid | Short Chain Fatty Acid | 0.21 | 0.0064 |
| sucrose | Carbohydrate | Disaccharides and Oligosaccharides | 0.21 | 0.0343 |
| N-formylphenylalanine | Amino Acid | Tyrosine Metabolism | 0.22 | 0.0200 |
| indoleacetylglutamine | Amino Acid | Tryptophan Metabolism | 0.23 | 0.0013 |
| 3-hydroxyhexanoylcarnitine (1) | Lipid | Fatty Acid Metabolism (Acyl Carnitine, Hydroxy) | 0.23 | 0.0137 |
| 3-methoxycatechol sulfate (1) | Xenobiotics | Benzoate Metabolism | 0.23 | 0.0331 |
| octadecadienedioate (C18:2-DC)* | Lipid | Fatty Acid, Dicarboxylate | 0.23 | 0.0405 |
| cyclo(leu-pro) | Peptide | Dipeptide | 0.24 | 0.0189 |
| 11beta-hydroxyandrosterone glucuronide | Lipid | Androgenic Steroids | 0.24 | 0.0193 |
| glucuronide of piperine metabolite C17H21NO3 (5)* | Xenobiotics | Food Component/Plant | 0.24 | 0.0759 |
| N-acetyl-aspartyl-glutamate (NAAG) | Amino Acid | Glutamate Metabolism | 0.26 | 0.0108 |
| N-linoleoyltaurine* | Lipid | Endocannabinoid | 0.27 | 0.0073 |
| glutamine conjugate of C6H10O2 (1)* | Partially Characterized Molecules | Partially Characterized Molecules | 0.27 | 0.0371 |
| 3-indoleglyoxylic acid | Xenobiotics | Food Component/Plant | 0.28 | 0.0016 |
| 4-hydroxyglutamate | Amino Acid | Glutamate Metabolism | 0.28 | 0.0051 |
| gamma-glutamylglutamate | Peptide | Gamma-glutamyl Amino Acid | 0.29 | 0.0029 |
| X-13866 | N/A | N/A | 0.29 | 0.0053 |
| methionine sulfoxide | Amino Acid | Methionine, Cysteine, SAM and Taurine Metabolism | 0.29 | 0.0075 |
| hydroquinone sulfate | Xenobiotics | Drug - Topical Agents | 0.29 | 0.0634 |
| linoleoyl ethanolamide | Lipid | Endocannabinoid | 0.30 | 0.0031 |
| isoleucylglycine | Peptide | Dipeptide | 0.30 | 0.0063 |
| cysteine sulfinic acid | Amino Acid | Methionine, Cysteine, SAM and Taurine Metabolism | 0.30 | 0.0108 |
| 3-carboxy-4-methyl-5-propyl-2-furanpropanoate (CMPF) | Lipid | Fatty Acid, Dicarboxylate | 0.30 | 0.0272 |
| gamma-glutamylvaline | Peptide | Gamma-glutamyl Amino Acid | 0.31 | 0.0030 |
| docosahexaenoate (DHA; 22:6n3) | Lipid | Long Chain Polyunsaturated Fatty Acid (n3 and n6) | 0.32 | 0.0040 |
| phenylacetylglutamate | Peptide | Acetylated Peptides | 0.32 | 0.0064 |
| eicosapentaenoate (EPA; 20:5n3) | Lipid | Long Chain Polyunsaturated Fatty Acid (n3 and n6) | 0.33 | 0.0154 |
| adipoylcarnitine (C6-DC) | Lipid | Fatty Acid Metabolism (Acyl Carnitine, Dicarboxylate) | 0.33 | 0.0154 |
| X-12101 | N/A | N/A | 0.33 | 0.0229 |
| 3-methoxycatechol sulfate (2) | Xenobiotics | Benzoate Metabolism | 0.33 | 0.0359 |
| Fibrinopeptide B (1-11)** | Peptide | Fibrinogen Cleavage Peptide | 0.33 | 0.0395 |
| gamma-glutamylphenylalanine | Peptide | Gamma-glutamyl Amino Acid | 0.34 | 0.0003 |
| arachidonate (20:4n6) | Lipid | Long Chain Polyunsaturated Fatty Acid (n3 and n6) | 0.35 | 0.0044 |
| (S)-3-hydroxybutyrylcarnitine | Lipid | Fatty Acid Metabolism (Acyl Carnitine, Hydroxy) | 0.35 | 0.0189 |
| homostachydrine* | Xenobiotics | Food Component/Plant | 0.35 | 0.0370 |
| X-12216 | N/A | N/A | 0.35 | 0.0798 |
| 3-carboxy-4-methyl-5-pentyl-2-furanpropionate (3-CMPFP)** | Lipid | Fatty Acid, Dicarboxylate | 0.36 | 0.0016 |
| arginine | Amino Acid | Urea cycle; Arginine and Proline Metabolism | 0.36 | 0.0024 |
| retinal | Cofactors and Vitamins | Vitamin A Metabolism | 0.37 | 0.0030 |
| metabolonic lactone sulfate | Partially Characterized Molecules | Partially Characterized Molecules | 0.37 | 0.0043 |
| cis-3,4-methyleneheptanoylcarnitine | Lipid | Fatty Acid Metabolism (Acyl Carnitine, Medium Chain) | 0.38 | 0.0042 |
| 1-stearoyl-GPC (18:0) | Lipid | Lysophospholipid | 0.38 | 0.0051 |
| 1-oleoylglycerol (18:1) | Lipid | Monoacylglycerol | 0.38 | 0.0103 |
| 3-hydroxyadipate | Lipid | Fatty Acid, Dicarboxylate | 0.38 | 0.0142 |
| glycochenodeoxycholate 3-sulfate | Lipid | Primary Bile Acid Metabolism | 0.38 | 0.0578 |
| 1-palmitoylglycerol (16:0) | Lipid | Monoacylglycerol | 0.39 | 0.0042 |
| 1-(1-enyl-palmitoyl)-GPC (P-16:0)* | Lipid | Lysoplasmalogen | 0.39 | 0.0060 |
| N-acetyl-isoputreanine | Amino Acid | Polyamine Metabolism | 0.39 | 0.0189 |
| 2-linoleoylglycerol (18:2) | Lipid | Monoacylglycerol | 0.39 | 0.0450 |
| orotate | Nucleotide | Pyrimidine Metabolism, Orotate containing | 0.39 | 0.0531 |
| glycerophosphorylcholine (GPC) | Lipid | Phospholipid Metabolism | 0.40 | 0.0009 |
| aspartate | Amino Acid | Alanine and Aspartate Metabolism | 0.40 | 0.0022 |
| 5-methyluridine (ribothymidine) | Nucleotide | Pyrimidine Metabolism, Uracil containing | 0.40 | 0.0404 |
| 2-palmitoleoyl-GPC (16:1)* | Lipid | Lysophospholipid | 0.40 | 0.0493 |
| N1-methylinosine | Nucleotide | Purine Metabolism, (Hypo)Xanthine/Inosine containing | 0.41 | 0.0020 |
| 1-(1-enyl-palmitoyl)-GPE (P-16:0)* | Lipid | Lysoplasmalogen | 0.41 | 0.0073 |
| 2-oleoylglycerol (18:1) | Lipid | Monoacylglycerol | 0.41 | 0.0212 |
| 1-stearoyl-GPI (18:0) | Lipid | Lysophospholipid | 0.41 | 0.0343 |
| pantothenate | Cofactors and Vitamins | Pantothenate and CoA Metabolism | 0.41 | 0.0532 |
| 1-stearoyl-GPG (18:0) | Lipid | Lysophospholipid | 0.41 | 0.0618 |

Table S5A. Plasma metabolites increased in obesity group pre- vs. post-sevelamer (*q<0.1*)

| **Metabolite** | **Family** | **Sub-Pathway** | **Fold change** | **q-value** |
| --- | --- | --- | --- | --- |
| 3-hydroxyhippurate sulfate | Xenobiotics | Benzoate Metabolism | 31.14 | 0.0650 |
| 4-ethylphenylsulfate | Xenobiotics | Benzoate Metabolism | 22.87 | 0.0542 |
| genistein sulfate* | Xenobiotics | Food Component/Plant | 21.80 | 0.0813 |
| umbelliferone sulfate | Xenobiotics | Food Component/Plant | 18.72 | 0.0663 |
| 3-hydroxyhippurate | Xenobiotics | Benzoate Metabolism | 14.33 | 0.0891 |
| trigonelline (N'-methylnicotinate) | Cofactors and Vitamins | Nicotinate and Nicotinamide Metabolism | 10.63 | 0.0076 |
| perfluorohexanesulfonic acid | Xenobiotics | Chemical | 8.63 | 0.0627 |
| taurocholate | Lipid | Primary Bile Acid Metabolism | 8.25 | 0.0822 |
| 3-hydroxypyridine sulfate | Xenobiotics | Chemical | 7.84 | 0.0019 |
| pentose acid* | Partially Characterized Molecules | Partially Characterized Molecules | 7.41 | 0.0224 |
| 3-methyl catechol sulfate (1) | Xenobiotics | Benzoate Metabolism | 7.36 | 0.0059 |
| heme | Cofactors and Vitamins | Hemoglobin and Porphyrin Metabolism | 7.20 | 0.0663 |
| 4-ethylcatechol sulfate | Xenobiotics | Benzoate Metabolism | 6.96 | 0.0047 |
| 3-acetylphenol sulfate | Xenobiotics | Chemical | 6.82 | 0.0999 |
| 5-acetylamino-6-formylamino-3-methyluracil | Xenobiotics | Xanthine Metabolism | 6.65 | 0.0089 |
| De(carboxymethoxy) cetirizine acetic acid | Xenobiotics | Drug - Respiratory | 5.86 | 0.0891 |
| N2-acetyl,N6-methyllysine | Amino Acid | Lysine Metabolism | 5.84 | 0.0316 |
| quinate | Xenobiotics | Food Component/Plant | 5.02 | 0.0019 |
| N-acetylcitrulline | Amino Acid | Urea cycle; Arginine and Proline Metabolism | 4.87 | 0.0343 |
| sedoheptulose | Carbohydrate | Pentose Metabolism | 4.75 | 0.0650 |
| tryptophan betaine | Amino Acid | Tryptophan Metabolism | 4.56 | 0.1120 |
| N2-acetyl,N6,N6-dimethyllysine | Amino Acid | Lysine Metabolism | 4.52 | 0.0468 |
| 5-acetylamino-6-amino-3-methyluracil | Xenobiotics | Xanthine Metabolism | 4.47 | 0.0365 |
| 1,3,7-trimethylurate | Xenobiotics | Xanthine Metabolism | 4.42 | 0.0274 |
| caffeine | Xenobiotics | Xanthine Metabolism | 4.34 | 0.0631 |
| stearoylcholine* | Lipid | Fatty Acid Metabolism (Acyl Choline) | 4.32 | 0.0468 |
| 1,7-dimethylurate | Xenobiotics | Xanthine Metabolism | 3.95 | 0.0511 |
| 2-oxoarginine* | Amino Acid | Urea cycle; Arginine and Proline Metabolism | 3.92 | 0.0343 |
| glutamine_degradant* | Partially Characterized Molecules | Partially Characterized Molecules | 3.85 | 0.0224 |
| picolinate | Amino Acid | Tryptophan Metabolism | 3.82 | 0.0518 |
| hippurate | Xenobiotics | Benzoate Metabolism | 3.81 | 0.0443 |
| S-methylcysteine sulfoxide | Amino Acid | Methionine, Cysteine, SAM and Taurine Metabolism | 3.78 | 0.0999 |
| 4-acetylphenol sulfate | Xenobiotics | Benzoate Metabolism | 3.63 | 0.0704 |
| pipecolate | Amino Acid | Lysine Metabolism | 3.51 | 0.0891 |
| guaiacol sulfate | Xenobiotics | Benzoate Metabolism | 3.49 | 0.0499 |
| 4-vinylphenol sulfate | Xenobiotics | Benzoate Metabolism | 3.48 | 0.0868 |
| paraxanthine | Xenobiotics | Xanthine Metabolism | 3.43 | 0.0617 |
| N-(2-furoyl)glycine | Xenobiotics | Food Component/Plant | 3.37 | 0.0252 |
| picolinoylglycine | Lipid | Fatty Acid Metabolism (Acyl Glycine) | 3.33 | 0.0019 |
| 1-methylxanthine | Xenobiotics | Xanthine Metabolism | 3.31 | 0.0544 |
| arachidonoylcholine | Lipid | Fatty Acid Metabolism (Acyl Choline) | 3.31 | 0.0555 |
| N-acetylglutamine | Amino Acid | Glutamate Metabolism | 3.25 | 0.0205 |
| desmethylnaproxen sulfate | Xenobiotics | Drug - Analgesics, Anesthetics | 3.25 | 0.0617 |
| 3-ethylcatechol sulfate (1) | Xenobiotics | Food Component/Plant | 3.23 | 0.0544 |
| 1-methylurate | Xenobiotics | Xanthine Metabolism | 3.15 | 0.0663 |
| ascorbic acid 3-sulfate* | Cofactors and Vitamins | Ascorbate and Aldarate Metabolism | 3.11 | 0.0059 |
| linoleoylcholine* | Lipid | Fatty Acid Metabolism (Acyl Choline) | 3.09 | 0.0835 |
| S-methylmethionine | Amino Acid | Methionine, Cysteine, SAM and Taurine Metabolism | 3.05 | 0.0813 |
| palmitoylcholine | Lipid | Fatty Acid Metabolism (Acyl Choline) | 2.98 | 0.0704 |
| 1-linoleoylglycerol (18:2) | Lipid | Monoacylglycerol | 2.97 | 0.0670 |
| theophylline | Xenobiotics | Xanthine Metabolism | 2.97 | 0.0712 |
| oleoylcholine | Lipid | Fatty Acid Metabolism (Acyl Choline) | 2.95 | 0.0750 |
| cyclo(leu-pro) | Peptide | Dipeptide | 2.78 | 0.0681 |
| 4-methylbenzenesulfonate | Xenobiotics | Chemical | 2.74 | 0.0693 |
| N6,N6-dimethyllysine | Amino Acid | Lysine Metabolism | 2.73 | 0.0544 |
| pyridoxate | Cofactors and Vitamins | Vitamin B6 Metabolism | 2.71 | 0.1036 |
| spermidine | Amino Acid | Polyamine Metabolism | 2.70 | 0.0542 |
| hydantoin-5-propionate | Amino Acid | Histidine Metabolism | 2.68 | 0.0208 |
| 2-oleoylglycerol (18:1) | Lipid | Monoacylglycerol | 2.68 | 0.0343 |
| 3-ureidopropionate | Nucleotide | Pyrimidine Metabolism, Uracil containing | 2.67 | 0.0215 |
| 2S,3R-dihydroxybutyrate | Lipid | Fatty Acid, Dihydroxy | 2.65 | 0.0627 |
| succinylcarnitine (C4-DC) | Energy | TCA Cycle | 2.64 | 0.0190 |
| hypotaurine | Amino Acid | Methionine, Cysteine, SAM and Taurine Metabolism | 2.62 | 0.0780 |
| N-acetylarginine | Amino Acid | Urea cycle; Arginine and Proline Metabolism | 2.57 | 0.0180 |
| formiminoglutamate | Amino Acid | Histidine Metabolism | 2.56 | 0.0260 |
| xanthurenate | Amino Acid | Tryptophan Metabolism | 2.56 | 0.0518 |
| oxindolylalanine | Amino Acid | Tryptophan Metabolism | 2.50 | 0.0468 |
| 1-stearoyl-2-arachidonoyl-GPS (18:0/20:4) | Lipid | Phosphatidylserine (PS) | 2.48 | 0.0609 |
| N2,N5-diacetylornithine | Amino Acid | Urea cycle; Arginine and Proline Metabolism | 2.46 | 0.0343 |
| (N(1) + N(8))-acetylspermidine | Amino Acid | Polyamine Metabolism | 2.42 | 0.0449 |
| catechol sulfate | Xenobiotics | Benzoate Metabolism | 2.40 | 0.0712 |
| isobutyrylcarnitine (C4) | Amino Acid | Leucine, Isoleucine and Valine Metabolism | 2.37 | 0.0544 |
| phenylacetylglutamine | Peptide | Acetylated Peptides | 2.35 | 0.0545 |
| glutamate, gamma-methyl ester | Amino Acid | Glutamate Metabolism | 2.35 | 0.0842 |
| homostachydrine* | Xenobiotics | Food Component/Plant | 2.34 | 0.0887 |
| 1-arachidonylglycerol (20:4) | Lipid | Monoacylglycerol | 2.33 | 0.0805 |
| 3-decenoylcarnitine | Lipid | Fatty Acid Metabolism (Acyl Carnitine, Monounsaturated) | 2.31 | 0.0215 |
| 1-lignoceroyl-GPC (24:0) | Lipid | Lysophospholipid | 2.31 | 0.0542 |
| 1-cerotoyl-GPC (26:0)* | Lipid | Lysophospholipid | 2.29 | 0.0663 |
| argininate* | Amino Acid | Urea cycle; Arginine and Proline Metabolism | 2.28 | 0.0545 |
| 3-methylglutarylcarnitine (2) | Amino Acid | Leucine, Isoleucine and Valine Metabolism | 2.27 | 0.0354 |
| N-acetyltaurine | Amino Acid | Methionine, Cysteine, SAM and Taurine Metabolism | 2.24 | 0.0479 |
| 4-hydroxyphenylacetylglutamine | Peptide | Acetylated Peptides | 2.24 | 0.0813 |
| 2-keto-3-deoxy-gluconate | Xenobiotics | Food Component/Plant | 2.18 | 0.0649 |
| 8-methoxykynurenate | Amino Acid | Tryptophan Metabolism | 2.17 | 0.0518 |
| 1-dihomo-linolenylglycerol (20:3) | Lipid | Monoacylglycerol | 2.16 | 0.0214 |
| naproxen | Xenobiotics | Drug - Analgesics, Anesthetics | 2.16 | 0.0650 |
| imidazole propionate | Amino Acid | Histidine Metabolism | 2.15 | 0.0719 |
| desmethylnaproxen | Xenobiotics | Drug - Analgesics, Anesthetics | 2.13 | 0.0704 |
| 3-amino-2-piperidone | Amino Acid | Urea cycle; Arginine and Proline Metabolism | 2.12 | 0.0807 |
| myo-inositol | Lipid | Inositol Metabolism | 2.03 | 0.0443 |
| methionine sulfone | Amino Acid | Methionine, Cysteine, SAM and Taurine Metabolism | 2.00 | 0.0518 |
| cysteine | Amino Acid | Methionine, Cysteine, SAM and Taurine Metabolism | 1.99 | 0.0215 |
| 3-formylindole | Xenobiotics | Food Component/Plant | 1.97 | 0.0518 |
| linoleoyl-arachidonoyl-glycerol (18:2/20:4) [2]* | Lipid | Diacylglycerol | 1.96 | 0.0542 |
| 5-(galactosylhydroxy)-L-lysine | Amino Acid | Lysine Metabolism | 1.95 | 0.0593 |
| N-acetylputrescine | Amino Acid | Polyamine Metabolism | 1.91 | 0.0365 |
| kynurenine | Amino Acid | Tryptophan Metabolism | 1.90 | 0.0343 |
| 1-palmitoyl-2-arachidonoyl-GPI (16:0/20:4)* | Lipid | Phosphatidylinositol (PI) | 1.90 | 0.0343 |
| 1-palmitoylglycerol (16:0) | Lipid | Monoacylglycerol | 1.88 | 0.0479 |
| octadecanedioate (C18-DC) | Lipid | Fatty Acid, Dicarboxylate | 1.88 | 0.0617 |
| asparagine | Amino Acid | Alanine and Aspartate Metabolism | 1.86 | 0.0343 |
| N-acetylhistidine | Amino Acid | Histidine Metabolism | 1.82 | 0.0576 |
| linoleoyl-arachidonoyl-glycerol (18:2/20:4) [1]* | Lipid | Diacylglycerol | 1.81 | 0.0717 |
| cortisol | Lipid | Corticosteroids | 1.79 | 0.0670 |
| 2-O-methylascorbic acid | Cofactors and Vitamins | Ascorbate and Aldarate Metabolism | 1.78 | 0.0365 |
| urea | Amino Acid | Urea cycle; Arginine and Proline Metabolism | 1.77 | 0.0627 |
| 1-myristoylglycerol (14:0) | Lipid | Monoacylglycerol | 1.77 | 0.0911 |
| cystine | Amino Acid | Methionine, Cysteine, SAM and Taurine Metabolism | 1.76 | 0.0836 |
| phenylpyruvate | Amino Acid | Phenylalanine Metabolism | 1.76 | 0.0891 |
| citrulline | Amino Acid | Urea cycle; Arginine and Proline Metabolism | 1.75 | 0.0147 |
| N,N-dimethyl-pro-pro | Peptide | Modified Peptides | 1.75 | 0.0603 |
| arabitol/xylitol | Carbohydrate | Pentose Metabolism | 1.74 | 0.0224 |
| 1-palmitoyl-2-linoleoyl-GPI (16:0/18:2) | Lipid | Phosphatidylinositol (PI) | 1.74 | 0.0468 |
| N1-methyladenosine | Nucleotide | Purine Metabolism, Adenine containing | 1.73 | 0.0518 |
| ornithine | Amino Acid | Urea cycle; Arginine and Proline Metabolism | 1.73 | 0.0693 |
| 4-methylcatechol sulfate | Xenobiotics | Benzoate Metabolism | 1.73 | 0.0943 |

Table S5B: Plasma metabolites decreased in obesity group pre- vs. post-sevelamer (*q<0.1*)

| **Metabolite** | **Family** | **Sub-Pathway** | **Fold change** | **q-value** |
| --- | --- | --- | --- | --- |
| 5-hydroxyindole sulfate | Amino Acid | Tryptophan Metabolism | 0.10 | 0.0741 |
| theanine | Xenobiotics | Food Component/Plant | 0.11 | 0.0544 |
| trans-3,4-methyleneheptanoate | Xenobiotics | Food Component/Plant | 0.32 | 0.0518 |
| andro steroid monosulfate C19H28O6S (1)* | Lipid | Androgenic Steroids | 0.44 | 0.0693 |
| glycine conjugate of C10H14O2 (1)* | Partially Characterized Molecules | Partially Characterized Molecules | 0.47 | 0.0763 |
| 10-undecenoate (11:1n1) | Lipid | Medium Chain Fatty Acid | 0.66 | 0.0767 |
| X-11478 | N/A | N/A | 0.70 | 0.0836 |

Table S6A: Plasma metabolites increased in obesity post-sevelamer vs. obesity post-placebo (*q<0.1*)

| **Metabolite** | **Family** | **Sub-Pathway** | **Fold change** | **q-value** |
| --- | --- | --- | --- | --- |
| caffeic acid sulfate | Xenobiotics | Food Component/Plant | 112.15 | 0.0805 |
| 1,2,3-benzenetriol sulfate (2) | Xenobiotics | Chemical | 39.84 | 0.0222 |
| 4-vinylcatechol sulfate | Xenobiotics | Benzoate Metabolism | 32.27 | 0.0406 |
| 4-ethylphenylsulfate | Xenobiotics | Benzoate Metabolism | 27.04 | 0.0216 |
| taurocholate | Lipid | Primary Bile Acid Metabolism | 22.29 | 0.0032 |
| genistein sulfate* | Xenobiotics | Food Component/Plant | 21.80 | 0.0286 |
| 3-hydroxyhippurate sulfate | Xenobiotics | Benzoate Metabolism | 17.14 | 0.0471 |
| trigonelline (N'-methylnicotinate) | Cofactors and Vitamins | Nicotinate and Nicotinamide Metabolism | 16.30 | 0.0178 |
| 9,10-DiHOME | Lipid | Fatty Acid, Dihydroxy | 14.88 | 0.0736 |
| umbelliferone sulfate | Xenobiotics | Food Component/Plant | 14.05 | 0.0247 |
| 3-methoxycatechol sulfate (1) | Xenobiotics | Benzoate Metabolism | 13.44 | 0.0056 |
| glycocholate | Lipid | Primary Bile Acid Metabolism | 12.96 | 0.0056 |
| 2-methoxyresorcinol sulfate | Xenobiotics | Chemical | 10.59 | 0.0512 |
| 4-acetylphenol sulfate | Xenobiotics | Benzoate Metabolism | 9.30 | 0.0213 |
| quinate | Xenobiotics | Food Component/Plant | 8.63 | 0.0378 |
| 3-hydroxypyridine sulfate | Xenobiotics | Chemical | 7.87 | 0.0235 |
| pyridoxate | Cofactors and Vitamins | Vitamin B6 Metabolism | 7.69 | 0.0032 |
| 4-ethylcatechol sulfate | Xenobiotics | Benzoate Metabolism | 7.69 | 0.0299 |
| 3-methoxycatechol sulfate (2) | Xenobiotics | Benzoate Metabolism | 7.56 | 0.0056 |
| 5alpha-androstan-3alpha,17beta-diol monosulfate (2) | Lipid | Androgenic Steroids | 7.52 | 0.0419 |
| octadecadienedioate (C18:2-DC)* | Lipid | Fatty Acid, Dicarboxylate | 7.37 | 0.0499 |
| N2-acetyl,N6,N6-dimethyllysine | Amino Acid | Lysine Metabolism | 6.85 | 0.0032 |
| 3-acetylphenol sulfate | Xenobiotics | Chemical | 6.64 | 0.0322 |
| pentose acid* | Partially Characterized Molecules | Partially Characterized Molecules | 6.49 | 0.0150 |
| 2,3-dihydroxy-2-methylbutyrate | Amino Acid | Leucine, Isoleucine and Valine Metabolism | 6.05 | 0.0446 |
| 1,2-dilinoleoyl-GPE (18:2/18:2)* | Lipid | Phosphatidylethanolamine (PE) | 5.98 | 0.0088 |
| De(carboxymethoxy) cetirizine acetic acid | Xenobiotics | Drug - Respiratory | 5.86 | 0.0333 |
| 11beta-hydroxyandrosterone glucuronide | Lipid | Androgenic Steroids | 5.46 | 0.0078 |
| 4-vinylphenol sulfate | Xenobiotics | Benzoate Metabolism | 5.22 | 0.0311 |
| glycoursodeoxycholic acid sulfate (1) | Lipid | Secondary Bile Acid Metabolism | 5.16 | 0.0868 |
| 4-methoxyphenol sulfate | Amino Acid | Tyrosine Metabolism | 5.01 | 0.0345 |
| pipecolate | Amino Acid | Lysine Metabolism | 4.86 | 0.0280 |
| taurochenodeoxycholate | Lipid | Primary Bile Acid Metabolism | 4.86 | 0.0726 |
| stearoylcholine* | Lipid | Fatty Acid Metabolism (Acyl Choline) | 4.77 | 0.0137 |
| sedoheptulose | Carbohydrate | Pentose Metabolism | 4.73 | 0.0174 |
| 4-guanidinobutanoate | Amino Acid | Guanidino and Acetamido Metabolism | 4.65 | 0.0091 |
| guaiacol sulfate | Xenobiotics | Benzoate Metabolism | 4.36 | 0.0110 |
| erythritol | Xenobiotics | Food Component/Plant | 4.20 | 0.0833 |
| tryptophan betaine | Amino Acid | Tryptophan Metabolism | 4.19 | 0.0526 |
| gamma-CEHC | Cofactors and Vitamins | Tocopherol Metabolism | 4.01 | 0.0103 |
| arachidonoylcholine | Lipid | Fatty Acid Metabolism (Acyl Choline) | 3.97 | 0.0214 |
| glucuronide of C10H18O2 (7)* | Partially Characterized Molecules | Partially Characterized Molecules | 3.92 | 0.0564 |
| behenoylcarnitine (C22)* | Lipid | Fatty Acid Metabolism (Acyl Carnitine, Long Chain Saturated) | 3.85 | 0.0361 |
| linoleoylcholine* | Lipid | Fatty Acid Metabolism (Acyl Choline) | 3.81 | 0.0198 |
| succinoyltaurine | Amino Acid | Methionine, Cysteine, SAM and Taurine Metabolism | 3.69 | 0.0032 |
| 4-hydroxyhippurate | Xenobiotics | Benzoate Metabolism | 3.63 | 0.0105 |
| indolepropionate | Amino Acid | Tryptophan Metabolism | 3.63 | 0.0805 |
| octadecenedioate (C18:1-DC) | Lipid | Fatty Acid, Dicarboxylate | 3.54 | 0.0339 |
| 1-linoleoylglycerol (18:2) | Lipid | Monoacylglycerol | 3.53 | 0.0083 |
| pantothenate | Cofactors and Vitamins | Pantothenate and CoA Metabolism | 3.52 | 0.0022 |
| N6-methyllysine | Amino Acid | Lysine Metabolism | 3.50 | 0.0121 |
| sucrose | Carbohydrate | Disaccharides and Oligosaccharides | 3.49 | 0.0209 |
| N2-acetyl,N6-methyllysine | Amino Acid | Lysine Metabolism | 3.49 | 0.0249 |
| palmitoylcholine | Lipid | Fatty Acid Metabolism (Acyl Choline) | 3.39 | 0.0234 |
| 4-imidazoleacetate | Amino Acid | Histidine Metabolism | 3.37 | 0.0428 |
| ascorbic acid 3-sulfate* | Cofactors and Vitamins | Ascorbate and Aldarate Metabolism | 3.35 | 0.0022 |
| 2-ketocaprylate | Amino Acid | Leucine, Isoleucine and Valine Metabolism | 3.30 | 0.0543 |
| 2-hydroxydecanoate | Lipid | Fatty Acid, Monohydroxy | 3.29 | 0.0484 |
| 2-aminooctanoate | Lipid | Fatty Acid, Amino | 3.28 | 0.0216 |
| 2-aminoheptanoate | Lipid | Fatty Acid, Amino | 3.27 | 0.0097 |
| desmethylnaproxen sulfate | Xenobiotics | Drug - Analgesics, Anesthetics | 3.25 | 0.0178 |
| 1-linoleoyl-GPG (18:2)* | Lipid | Lysophospholipid | 3.20 | 0.0035 |
| catechol sulfate | Xenobiotics | Benzoate Metabolism | 3.19 | 0.0162 |
| glutamate, gamma-methyl ester | Amino Acid | Glutamate Metabolism | 3.12 | 0.0072 |
| N-(2-furoyl)glycine | Xenobiotics | Food Component/Plant | 3.12 | 0.0143 |
| 1-linoleoyl-GPE (18:2)* | Lipid | Lysophospholipid | 3.11 | 0.0091 |
| 3-methyl catechol sulfate (1) | Xenobiotics | Benzoate Metabolism | 3.09 | 0.0622 |
| N-acetyl-2-aminooctanoate* | Lipid | Fatty Acid, Amino | 3.07 | 0.0087 |
| N6,N6-dimethyllysine | Amino Acid | Lysine Metabolism | 3.07 | 0.0091 |
| 1-lignoceroyl-GPC (24:0) | Lipid | Lysophospholipid | 3.05 | 0.0062 |
| oleoylcholine | Lipid | Fatty Acid Metabolism (Acyl Choline) | 3.04 | 0.0412 |
| linoleoyl-linoleoyl-glycerol (18:2/18:2) [2]* | Lipid | Diacylglycerol | 3.03 | 0.0078 |
| tetrahydrocortisone glucuronide (5) | Lipid | Corticosteroids | 3.00 | 0.0119 |
| 5-acetylamino-6-amino-3-methyluracil | Xenobiotics | Xanthine Metabolism | 2.99 | 0.0232 |
| glycochenodeoxycholate 3-sulfate | Lipid | Primary Bile Acid Metabolism | 2.99 | 0.0379 |
| homostachydrine* | Xenobiotics | Food Component/Plant | 2.95 | 0.0222 |
| 2-linoleoylglycerol (18:2) | Lipid | Monoacylglycerol | 2.95 | 0.0235 |
| adipoylcarnitine (C6-DC) | Lipid | Fatty Acid Metabolism (Acyl Carnitine, Dicarboxylate) | 2.94 | 0.0220 |
| delta-CEHC | Cofactors and Vitamins | Tocopherol Metabolism | 2.93 | 0.0131 |
| 1-linolenoylglycerol (18:3) | Lipid | Monoacylglycerol | 2.92 | 0.0051 |
| 3-methylglutarylcarnitine (2) | Amino Acid | Leucine, Isoleucine and Valine Metabolism | 2.92 | 0.0234 |
| (N(1) + N(8))-acetylspermidine | Amino Acid | Polyamine Metabolism | 2.90 | 0.0061 |

Table S6B: Plasma metabolites decreased in obesity post-sevelamer vs. obesity post-placebo (*q<0.1*)

| **Metabolite** | **Family** | **Sub-Pathway** | **Fold change** | **q-value** |
| --- | --- | --- | --- | --- |
| hydroxybupropion | Xenobiotics | Drug - Psychoactive | 0.01 | 0.0468 |
| pregnanediol-3-glucuronide | Lipid | Progestin Steroids | 0.18 | 0.0817 |
| gamma-glutamylalanine | Peptide | Gamma-glutamyl Amino Acid | 0.48 | 0.0234 |
| hyocholate | Lipid | Secondary Bile Acid Metabolism | 0.51 | 0.0633 |
| piperine | Xenobiotics | Food Component/Plant | 0.53 | 0.0445 |

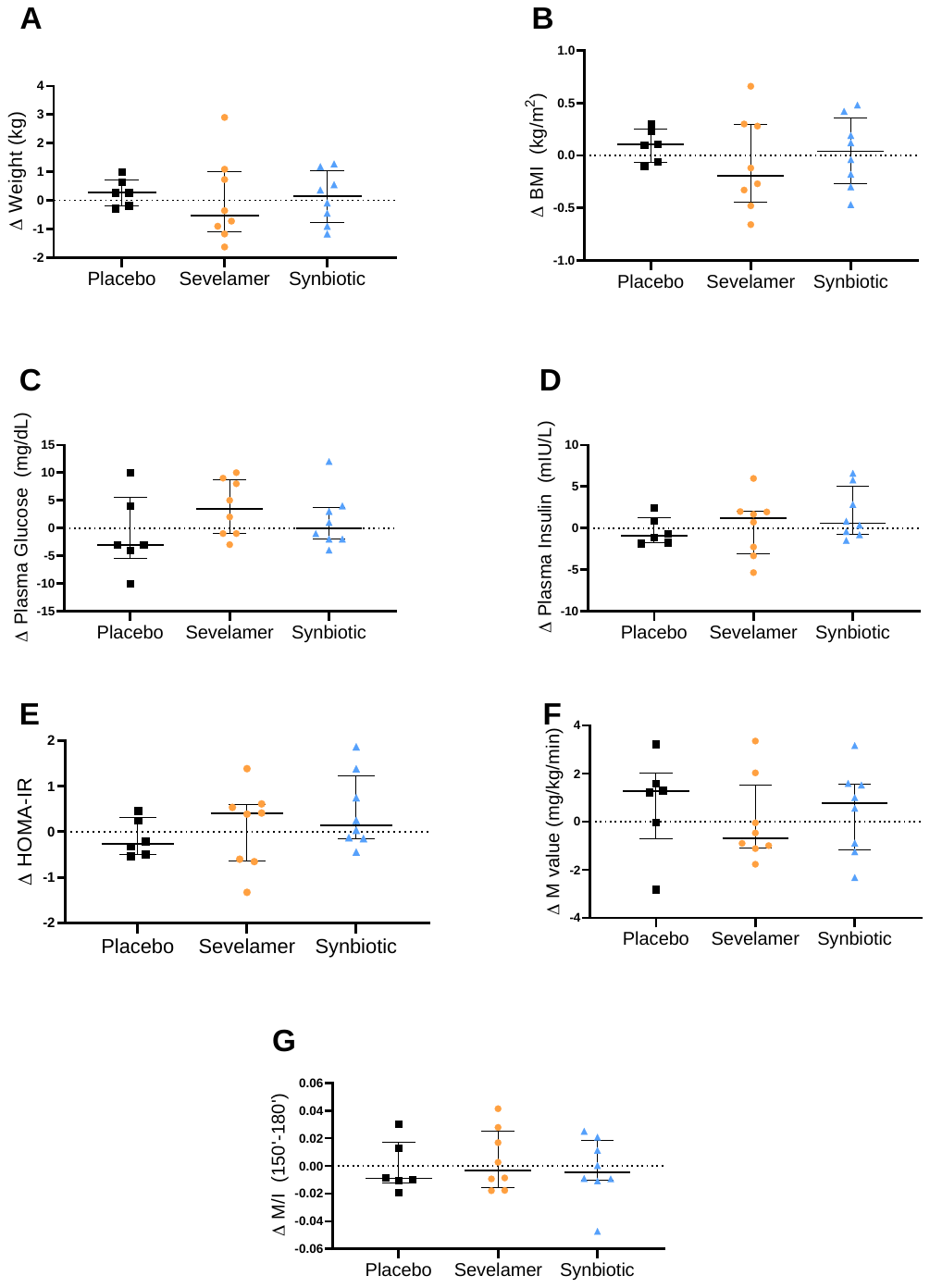

**Figure S1. Effect of placebo, sevelamer and synbiotic on glucose metabolism outcomes in lean subjects.** (A) Body weight, (B) BMI, (C) fasting plasma glucose, (D) fasting plasma insulin, (E) HOMA-IR, (F) M value, and (G) M/I in lean subjects. Within-intervention effects were analyzed using paired t-test; *p<0.05. Between intervention effects analyzed using generalized estimating equations; *p<0.025. Data are means ± SEM.

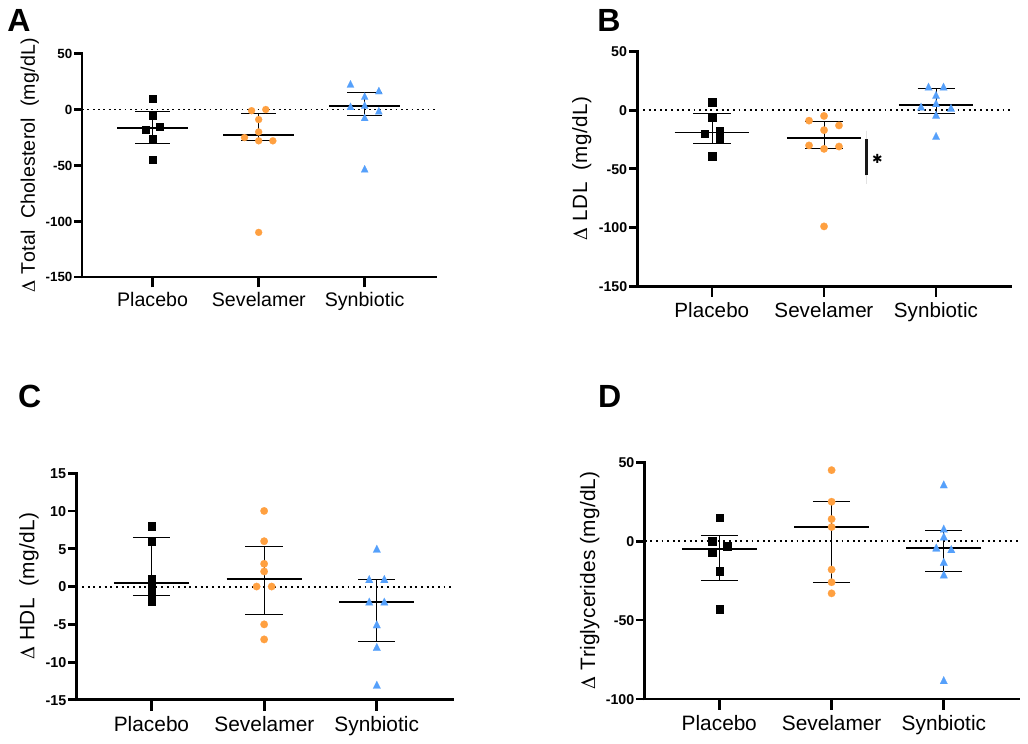

**Figure S2. Effect of placebo, sevelamer and synbiotic on serum lipid profile in lean subjects.** (A) Fasting serum total cholesterol, (B) LDL cholesterol, (C) HDL cholesterol and (D) triglycerides. Within-intervention effects were analyzed using paired t-test; *p<0.05. Between intervention effects analyzed using generalized estimating equations; *p<0.025. Data are means ± SEM.

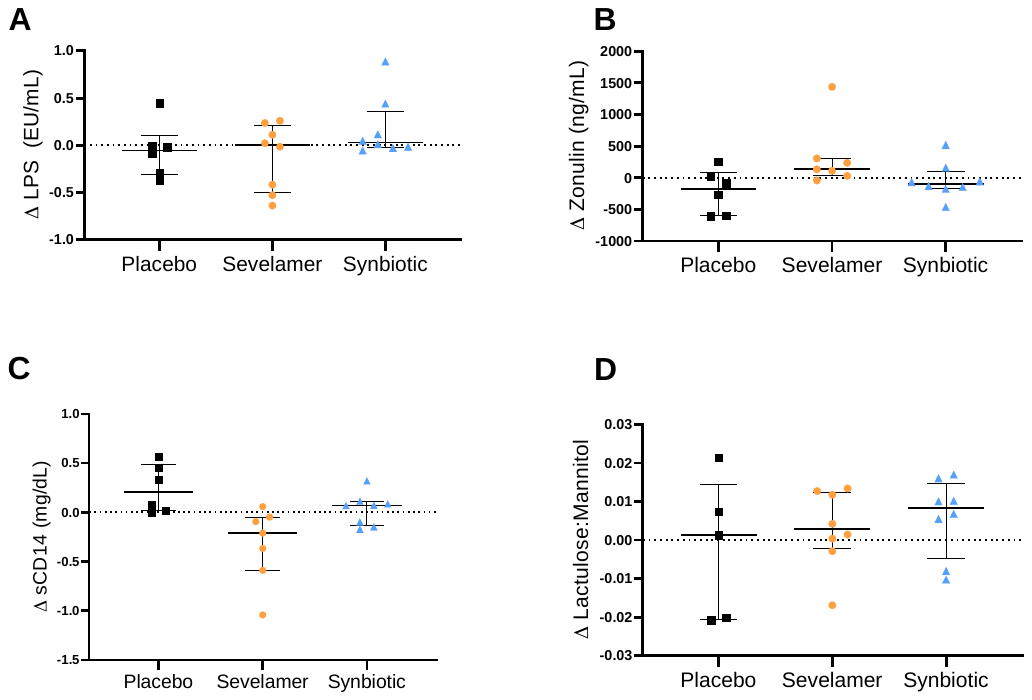

**Figure S3. Effect of placebo, sevelamer and synbiotic on measures of metabolic endotoxemia and intestinal permeability in lean subjects.** (A) fasting plasma LPS, (B) zonulin, (C) sCD14, and (D) lactulose:mannitol. Within-intervention effects were analyzed using paired t-test; *p<0.05. Between intervention effects analyzed using generalized estimating equations; *p<0.025. Data are means ± SEM.
